# Both ageing and frailty status impact vaccine-induced transcriptomic profiles and subsequent humoral immunity: results from the VITAL cohort

**DOI:** 10.64898/2026.08.26.26361408

**Authors:** Manas Joshi, Christophe Carré, Alper Çevirgel, Elske Bijvank, Martine Chabaud-Riou, Virginie Courtois, Emilie Chautard, Daniel Larocque, Wivine Burny, Lisa Beckers, Anne-Marie Buisman, Nynke Rots, Marieke van der Heiden, Josine van Beek, Yannick van Sleen, Debbie van Baarle

## Abstract

Vaccine responses vary across individuals due to differences in ageing and health status. Using transcriptomic profiling, we analyzed early gene expression profiles after influenza (QIV) followed by pneumococcal (PCV13) vaccination in 148 participants spanning young, middle-aged, and older adults. The two vaccines induced distinct immune signatures: QIV elicited innate and interferon immune activation, while PCV13 triggered inflammation-based responses. Older adults showed weaker but similar transcriptomic profiles compared to young adults. Among older adults, frailty, in addition to age, was strongly associated with reduced innate responses. In addition, we identified associations between early-stage transcriptomic profiles and later-stage antibody responses for QIV; however, no such associations were observed for PCV13. Importantly, observed group differences arose not from altered immune modules but from differences in the magnitude of gene expression, paving the way for immune-boosting interventions to enhance early gene expression in at-risk populations.

## INTRODUCTION

Vaccination has been shown to be an effective intervention strategy for priming the immune system in combating pathogens, thereby acting as a potent tool for healthy ageing. Over the past century, vaccines have been instrumental in lowering the morbidity and mortality rates associated with various infectious diseases (*1–3*), reducing the burden on healthcare facilities. Overall, vaccine efficacy is significantly influenced by chronological age, and functional deterioration of the immune system with age – immunosenescence – has been suggested to be a main contributor to this declined response (*4–6*). Immunosenescence captures a variety of age-associated functional changes of the immune system, which impair its ability to combat pathogens and respond to vaccines (*7–9*).

Chronological ageing impacts individuals differently (*10*), with some following a healthy ageing trajectory and others displaying an accelerated accumulation of comorbidities leading to a state of frailty, which is associated with various adverse clinical outcomes (*11,12*). These different ageing trajectories, but also other individual-specific factors (e.g., sex, immunization history, exposure to pathogens, etc.), could influence the effectiveness of the immune system in responding to infections and vaccines. These immune differences lead to distinct post-vaccination responses within a heterogeneous population. Characterizing these different immune response profiles, especially low response profiles, is important to identify individuals in need of alternative vaccination strategies to enhance immune protection by vaccination.

Transcriptomics enables deeper insights into characterizing the vaccine responses through differential gene expression patterns, thereby uncovering the molecular mechanisms underlying the vaccine response profiles. In a nutshell, using the relative abundance or reduction in transcripts post-vaccination in comparison to pre-vaccination, transcriptomics elucidates the genes and the associated biological pathways impacted by vaccination. Differential gene expression analyses have identified key vaccine-driven immunological processes, including innate and humoral responses, revealing pathways associated with immune responses (*13–17*). To exemplify, using transcriptomics approaches, Carre *et al*. 2021 (*18*) elucidated key molecular mechanisms underlying immunosenescence, such as dysregulated endoplasmic reticulum stress responses and altered bile acid metabolism, that likely contribute to diminished vaccine efficacy in older adults.

While transcriptomics has been used to explore post-vaccination innate and adaptive immune responses, few studies have compared responses to different vaccines within the same individual, and none have done so across age groups within the same cohort (but see (*18–20*) for other cohorts). To address this knowledge gap, this study characterized and compared gene expression signatures induced by a booster quadrivalent inactivated influenza vaccine (QIV) and a primary 13-valent pneumococcal-conjugate vaccine (PCV13) in the blood of participants within the VITAL (Vaccines and Infectious Diseases in the Ageing Population) cohort (*21,22*). VITAL is a unique healthy ageing cohort where all participants were consecutively vaccinated with two different vaccines (QIV and PCV-13). Through its unique design, this study aimed to identify factors underlying vaccine responsiveness across multiple vaccines. One of the key recruitment criteria was previous vaccination against influenza, and no prior pneumococcal vaccination. The primary outcomes of this vaccination trial revealed that older adults had weaker humoral immune responses as well as a lower persistence of humoral immunity compared to younger adults, although individual responses to different vaccines varied substantially (*22*).

In this exploratory study, we aimed to unravel the early transcriptome profiles induced after QIV followed by PCV13 vaccination in the same individuals. To this end, we compared the gene expression profiles at 1,2 and 7 days post-vaccination between the QIV and PCV13 vaccines using whole blood samples. Given the heterogeneous antibody profiles observed (*22*), we also studied the effect of chronological age, frailty and sex on the post-vaccination transcriptome profiles for both vaccines. Additionally, using the humoral response data, we assessed the association between the early post-vaccination transcriptome profiles and the later-stage antibody levels.

## RESULTS

### Study population

The VITAL cohort consists of 326 participants (*22*), of which the transcriptomics (TC) sub-cohort was set up comprising 148 individuals shortlisted based on their antibody data availability for both the QIV and the PCV13 vaccines and sampling day alignments (see <u>Materials and Methods</u>). Based on the original cohort setup, these individuals were categorized into three age categories: young adults (YA, aged 25-49 years, n = 32), middle-aged adults (MA, aged 50-64 years, n = 60), and older adults (OA, aged 65 years and older, n = 56). Transcriptome data were collected and analyzed at the following timepoints: pre-vaccination (day 0) and post-vaccination (day 1 or 2, and day 7). All differential expression patterns were determined using the pre-vaccination timepoint as the baseline. The two vaccines had their own independent baselines, and all analyses were performed using these baselines as reference.

The baseline characteristics of the TC sub-cohort are listed in [Table 1] and were overall similar to the full cohort (*22*). Distributions of the listed parameters were found to be comparable between the TC sub-cohort and the rest of the cohort (Non-TC) [Supplementary Figure 8, BH adjusted p-value > 0.05]. Our findings from the primary study (*22*) indicated that QIV-induced antibody titers decreased progressively with age. In addition, PCV13 responses were characterized by broader serotype coverage in young adults (YA) relative to middle-aged (MA) and older adults (OA).

**Table 1:**
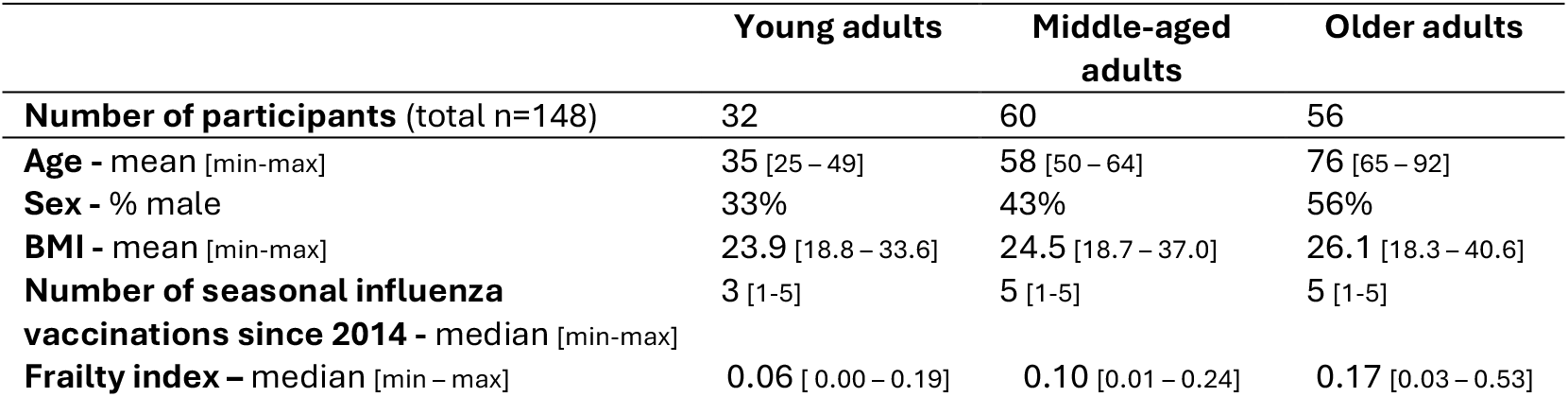
Demographic and clinical characteristics of the transcriptome sub-cohort. Values are presented as medians, with ranges (minimum–maximum) indicated in square brackets.

### QIV and PCV13 induce activation of different blood transcriptome modules consistent with their composition and mechanism of action

We analyzed the post-vaccination transcriptome profiles (PTPs) predominantly through a blood transcriptome module (BTM)-based approach as described in Chaussabel et al., 2008 and Li et al., 2014 (*23*,*24*). To gain a comprehensive understanding of biological responses impacted by vaccination, we aggregated related BTMs into ‘super-modules’ and scored them using both the proportion and magnitude of differentially expressed genes within each module (see Materials and Methods). Here, super-modules are collections of related BTMs that are reflective of specific immune processes. Due to logistical constraints, individuals were either analysed at Day 1 (n=86) or Day 2 (n=62), post-vaccination (see <u>Materials and Methods</u>). For Day 7 we could analyse all individuals (n=148).

Overall, PCV13 triggered more differentially expressed genes (DEGs) compared to QIV, and both vaccines showed a relatively higher DEG count on day 1 [Supplementary Table 10]. We noted subtle global-level differences in the gene expression profiles across the three post-vaccination timepoints for the two vaccines [Figure 1(a)]. The top five upregulated and downregulated DEGs differed between the two vaccines at day 1 [Figure 1(b)]. PCV13 showed prominent upregulation of inflammation-associated genes (CASP5 and PROK2), whereas QIV showed expression of innate immunity-related genes (ETV7, GBP1, and GBP5). When comparing PTPs originating from the QIV (inactivated antiviral booster) vaccine and the PCV13 (conjugate antibacterial primary) vaccine, we noted clear differences [Figure 1(c)]. At day 1, QIV induced upregulation of interferon and innate immunity, and antigen presentation cell-based (APCs) modules. In contrast, PCV13 induced upregulation of inflammation and APC modules, in addition to downregulation of the T cell modules. On day 7, we noted comparatively higher upregulation of the B cell-based module (LI.M156.1: plasma cells, immunoglobulins) for PCV13 as compared to QIV.

**Figure 1:**
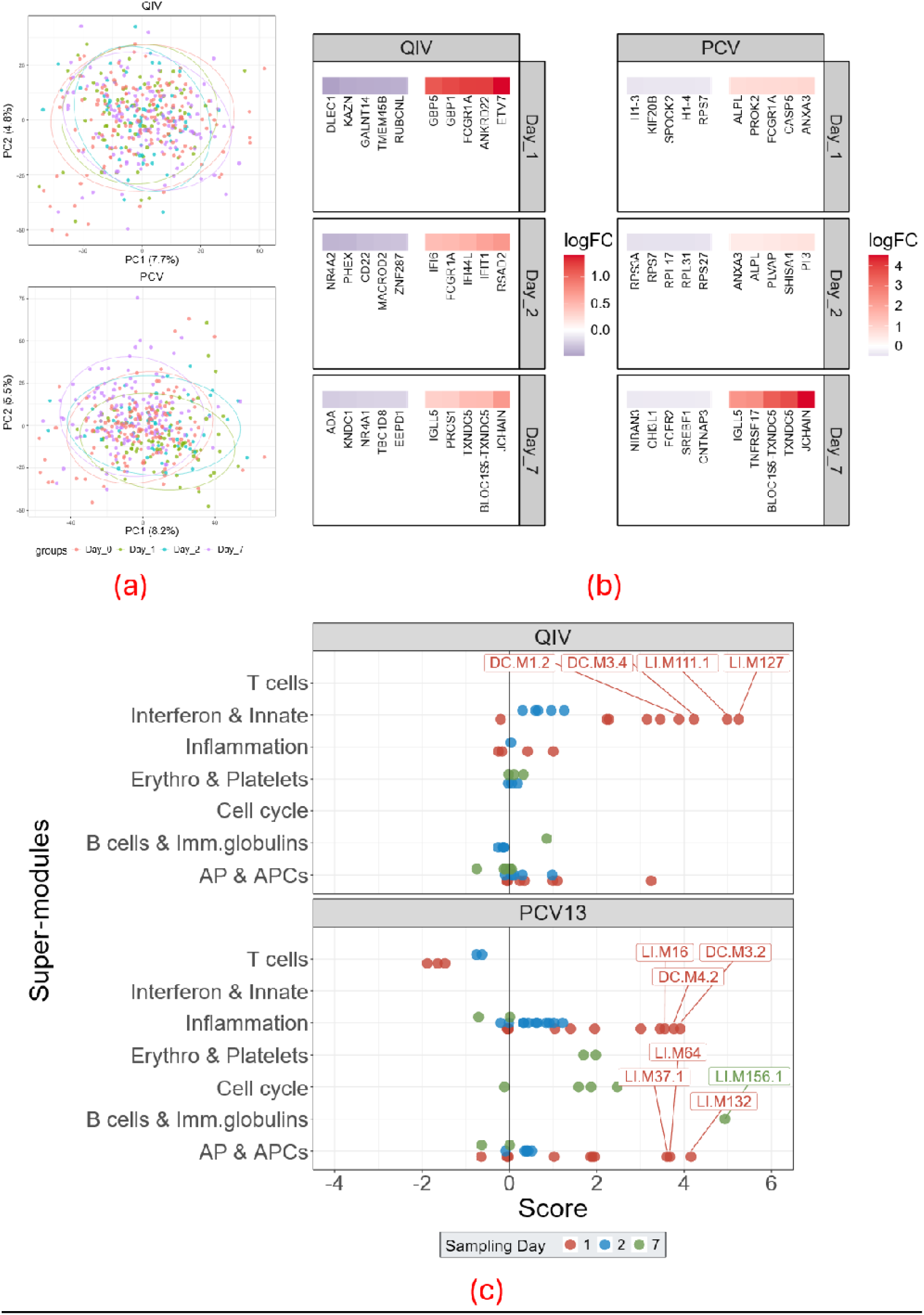
Overall post-vaccination vaccine gene expression changes for the two vaccines; (a) Principal component analysis (PCA) plots illustrating global gene expression patterns across post-vaccination timepoints; (b) Heatmaps showing the top five significantly upregulated and downregulated genes (p-value < 0.05) across all post-vaccination timepoints; (c) Post-vaccination transcriptome profile (PTP) differences for QIV (top panel) and PCV13 (bottom panel). The X-axis represents module scores and the Y-axis shows grouped modules (super-modules). Each dot corresponds to a single blood transcriptome module (BTM), colored by post-vaccination timepoint (day 1 – red, day 2 – blue, day 7 – green). Negative scores indicate enrichment of downregulated genes, while positive scores indicate enrichment of upregulated genes. Modules with scores >3.5 are annotated using Li et al., 2014 (24) and Chaussabel et al., 2008 (23) nomenclature [see Supplementary Table 11]. Scores are indicative of the proportion of DEG and their respective fold changes (negative score – downregulated genes; positive score – upregulated genes) per module

### Differences in the PTPs across age categories and sexes

Due to the age-associated functional deterioration of the immune system, differences in gene expression after vaccination would be expected with age. We note subtle changes in the overall gene expression profiles for the two vaccines across all age groups at the post-vaccination timepoints [Supplementary Figure 9]. Interestingly, for both vaccines, all age groups also showed up- and downregulation of similar blood transcriptome modules (BTMs) in their PTPs, predominantly at day 1 post-vaccination [Figure 2]. However, the expression levels differed between age groups. Specifically, for QIV, innate immunity and interferon gene modules were significantly more upregulated in the YA as compared to OA [Supplementary Table 1, Benjamini-Hochberg (BH) adjusted p-value < 0.05]. Similarly, for PCV13, the inflammation modules were significantly more upregulated in YA compared to OA [Supplementary Table 1, BH adjusted p-value < 0.05]. YA also showed higher DEGs as compared to OA at day 1 for both vaccines [Supplementary Table 10]. Additionally, we noted downregulation of T cell modules exclusively for the YA group for both vaccines.

**Figure 2:**
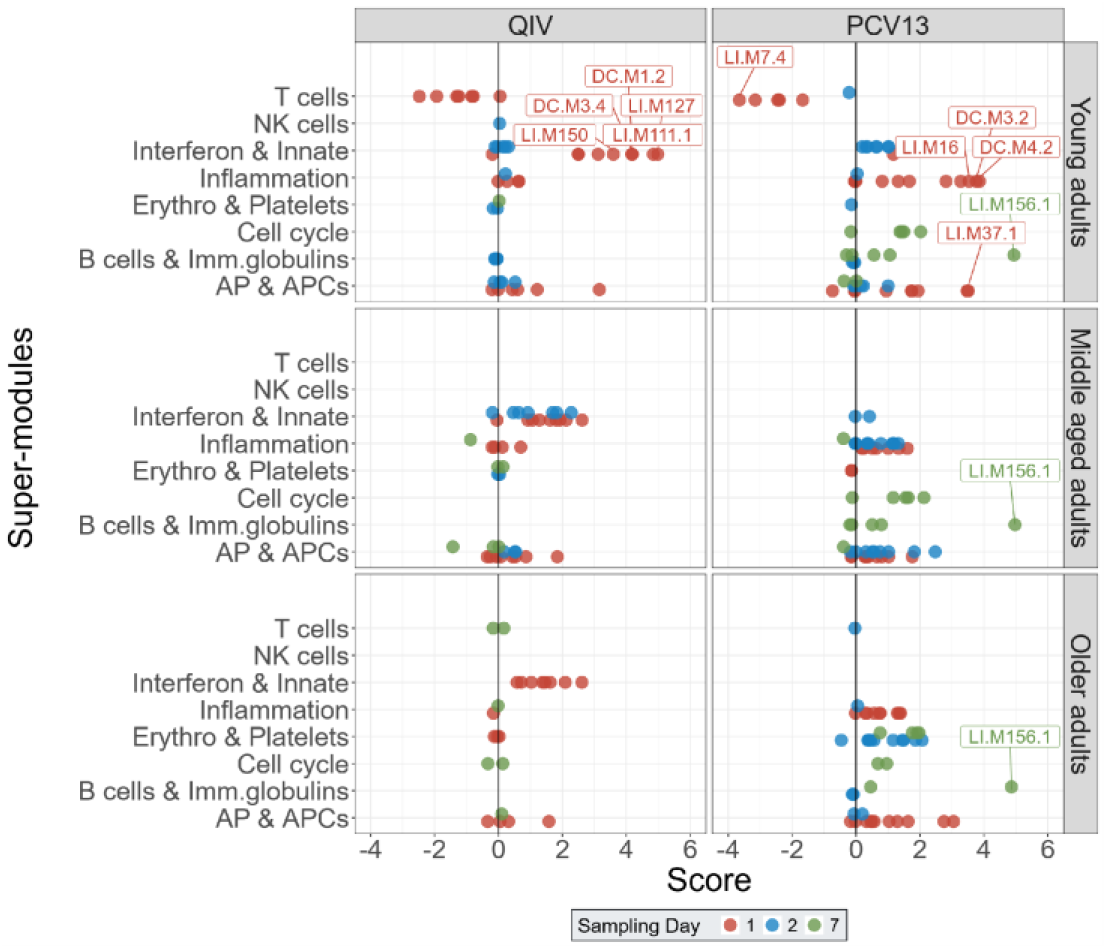
Post-vaccination transcriptome profiles (PTPs) for the two vaccines split by the three age categories. Every dot corresponds to a single BTM, and its corresponding score reflects an excess of significantly upregulated (positive score) or downregulated (negative score) genes. The colours of the dots are reflective of the post-vaccination sampling time point. Modules with scores higher than 3.5 are annotated with the Li et al., 2014 (24) and Chaussabel et al., 2008 (23) nomenclature [see Supplementary Table 11]. Scores are indicative of the proportion of DEG and their respective fold changes (negative score – downregulated genes; positive score – upregulated genes) per module

As we also observed differences in the antibody levels between males and females to QIV in the primary cohort (*22*), we also studied sex-specific PTPs in response to both vaccines. Interestingly, we noted no significant differences in the PTPs for the two sexes in response to PCV13 [Figure 3]. However, in the case of QIV, at day 1, the innate and interferon, and inflammation-based transcriptome modules were significantly more upregulated in females than males [Supplementary Table 2, BH adjusted p-value < 0.05].

**Figure 3:**
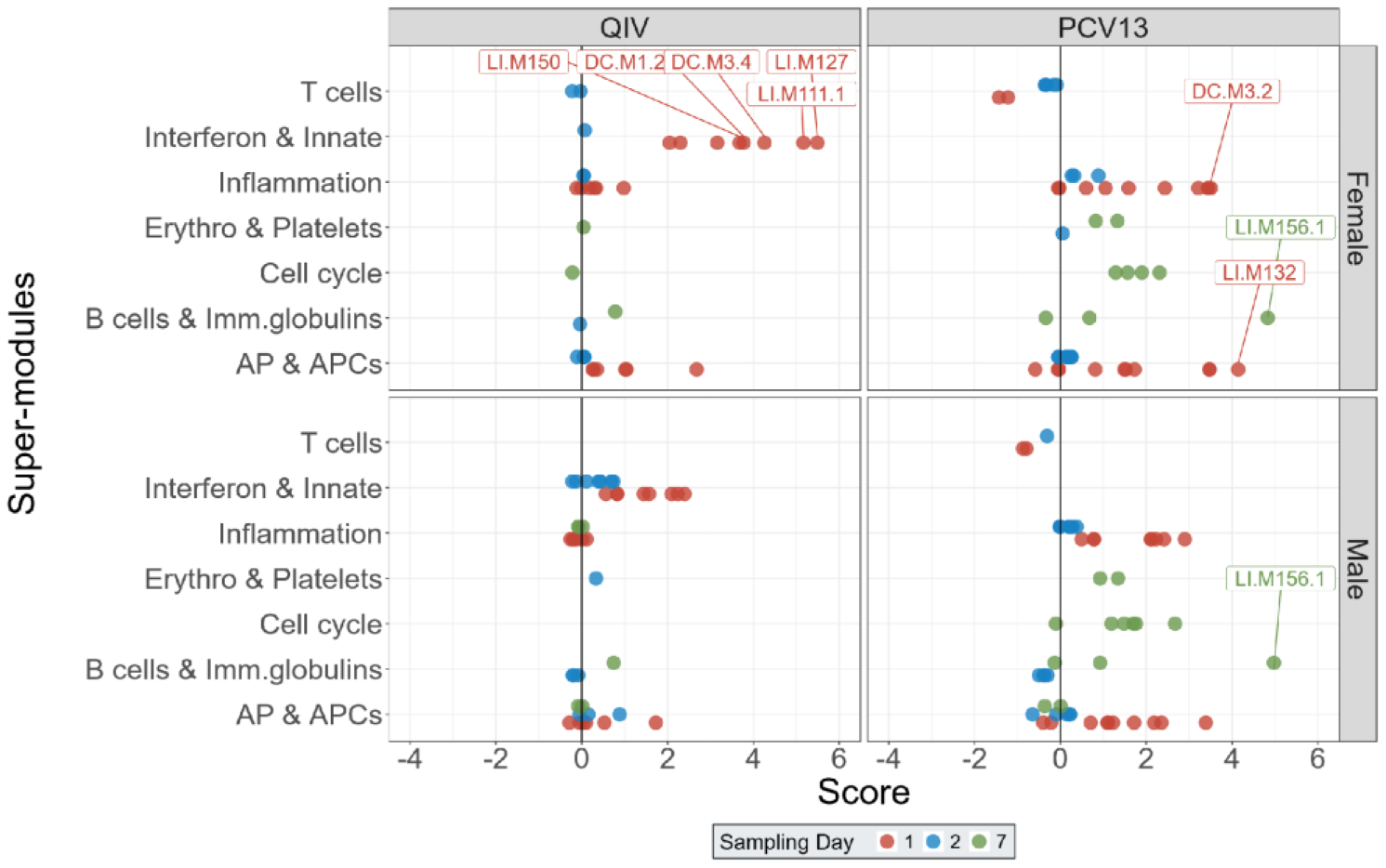
Sex-specific post-vaccination transcriptome profiles (PTPs) split for the two vaccines. The individual BTMs are indicated as dots and grouped into super-modules. Colors of the BTMs are indicative of the sampling days, and the score is indicative of the overall upregulation (positive) or downregulation (negative) of genes contained within them. Modules with scores higher than 3.5 are annotated with the Li *et al*., 2014 (*24*) and Chaussabel *et al*., 2008 (*23*) nomenclature [see Supplementary Table 11]. Scores are indicative of the proportion of DEG and their respective fold changes (negative score – downregulated genes; positive score – upregulated genes) per module

### Age-corrected non-frail groups showcase elevated levels of transcriptome module activation

Next, we assessed the impact of frailty status on the PTPs. Given the established link between immune function and overall well-being, the frailty index helps in quantifying immune functional status, serving as a useful functional proxy for biological ageing. Using the frailty index derived from 31 health deficits and calculated for each individual in the cohort separately (*11*), we performed local regression analysis on the age and frailty parameters in the older age groups (MA and OA), where frailty is most relevant. To identify effects of frailty relative to an individual’s chronological age, we developed an age-corrected frailty grouping strategy by using the regression line and its 95% confidence interval [Figure 4a, blue line and grey shaded area] to make an age-specific regression fit. Individuals falling below the age–frailty regression line were classified as non-frail, indicating lower frailty index values than expected for their age [Figure 4(a), n=44, age-corrected non–frail, green data points]. Conversely, individuals above the regression line were classified as frail [Figure 4(a), n=33, age-corrected frail, orange data points]. Besides the distribution of the frailty scores and the number of medications (which is incorporated in frailty score calculations), the two categories did not differ significantly in demographic (age and sex) or other clinical variables (BMI, a component of the frailty index) [Supplementary Table 3].

**Figure 4:**
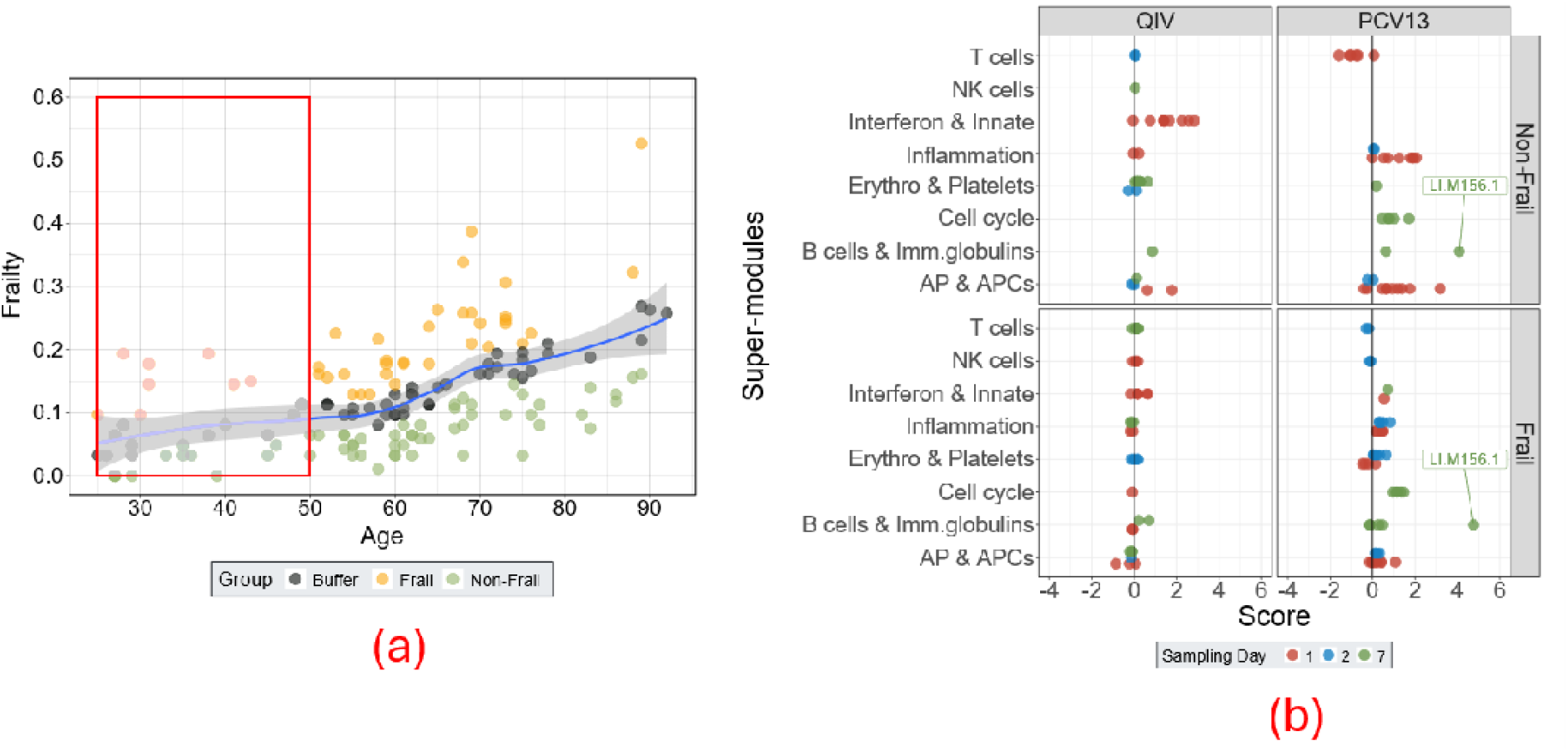
Gene expression changes associated with age-corrected frailty groups; (a) Local regression between frailty and age parameters. Each dot represents an individual. Based on the regression line, individuals were classified as frail (above the fit) or non-frail (below). Young adults (YA) were omitted from this analysis, indicated by the faded red box; only middle-aged (MA) and older adults (OA) were included; (b) The PTPs for the two vaccines split into the age-corrected frailty groups. The individual BTMs are indicated as a single dot and are coloured based on the sampling timepoint post-vaccination. The modules are scored based on the enrichment of genes significantly upregulated (positive score) or downregulated (negative score). Modules with scores higher than 3.5 are annotated with the Li et al., 2014 (24) and Chaussabel et al., 2008 (23) nomenclature [see Supplementary Table 11]. Scores are indicative of the proportion of DEG and their respective fold changes (negative score – downregulated genes; positive score – upregulated genes) per module

We noted subtle changes in the overall gene expression profiles for the two groups [Supplementary Figure 9]. Next, we compared the PTPs of the two categories [Figure 4(b)]. Interestingly, for QIV, the BTM-based interferon and innate modules were significantly more upregulated in non-frail than in frail groups [Supplementary Table 4, BH adjusted p-value < 0.05]. In the case of PCV13, the inflammation modules were significantly more upregulated, and T cell-related modules were more downregulated in non-frail compared to frail groups [Supplementary Table 4, BH adjusted p-value < 0.05], mirroring the pattern observed in YA. Additionally, the non-frail group showed relatively higher DEGs at day 1 for both vaccines [Supplementary Table 10].

Interestingly, despite having similar frailty indices, older individuals who were relatively healthy (Non-frail group, Median frailty index – 0.07, Supplementary Table 3) exhibited relatively weaker transcriptomic profiles than YA (Median frailty index – 0.06, Table 1) [Supplementary Figure 5]. In contrast, older individuals who were relatively more frail (Frail group, Median frailty index – 0.21, Supplementary Table 3) than expected for their age showed markedly diminished transcriptome profiles, lower even than the average for the older (Median frailty index – 0.17, Table 1) cohort [Supplementary Figure 6].

### Vaccine-specific associations between early-stage PTPs and late-stage antibody titers

To understand the relation between early gene expression and subsequent humoral immune response dynamics, we next focused on identifying associations between the early-stage gene expression profile and the individual-specific elicited antibody titers. To this end, we compared post-vaccination transcriptome profiles (PTPs) between the individual-specific antibody response (based on 28-day post-vaccination antibody titers) stratification quartiles outlined in the primary cohort (*22*). In brief, individuals were assigned a quartile-based score indicating their relative antibody titer levels. A score of 1 corresponded to the lowest titers, a score of 4 to the highest titers, and scores of 2 and 3 to intermediate titer levels. The age group distribution in the quartiles is depicted in [Supplementary Figure 1].

Here, we focused our analyses on the lowest (score 1) and highest (score 4) titer groups. Low and high antibody responders again showed PTP dynamics of similar modules, albeit with varying levels [Figure 5]. Overall, high responders showcased higher DEGs than low responders for both vaccines [Supplementary Table 10]. For QIV, at day 1, high antibody responders showed significantly higher upregulation of interferon and innate immunity modules [Supplementary Table 5, BH adjusted p-value < 0.05]. Interestingly, the innate immunity-associated gene expression patterns were also present at day 2 exclusively for high responders. For PCV13, we observed that high and low responders showed comparable and strong upregulation of inflammation and APC-related modules, but at different time points: day 1 for high responders and day 2 for low responders.

**Figure 5:**
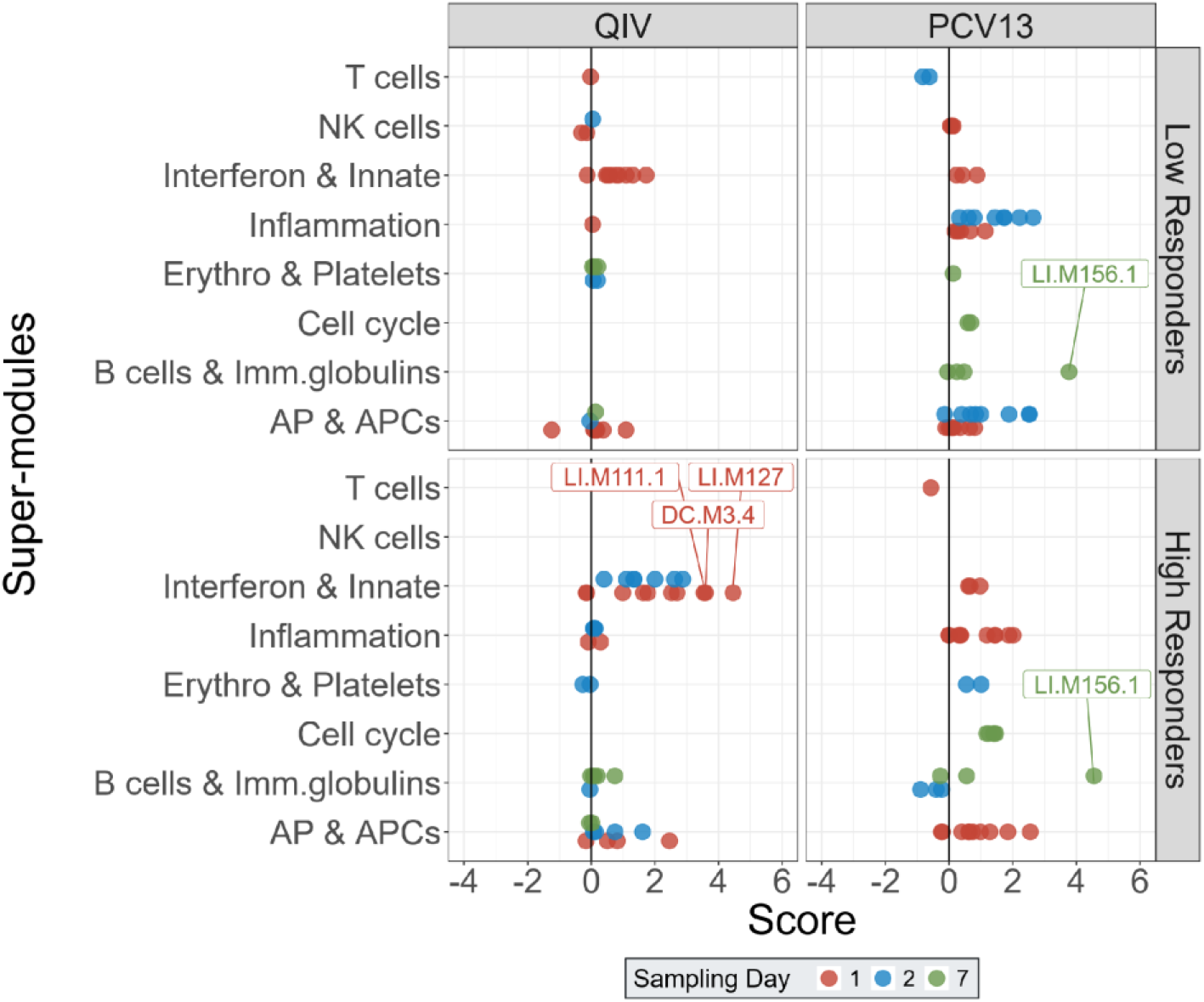
The PTPs for the two vaccines split into the high (Responder 4) and low (Responder 1) responder categories. The individual BTMs are indicated as a single dot and are coloured based on the sampling timepoint post-vaccination. The modules are scored based on the enrichment of genes significantly upregulated (positive score) or downregulated (negative score). Modules with scores higher than 3.5 are annotated with the Li *et al*., 2014 (*24*) and Chaussabel *et al*., 2008 (*23*) nomenclature [see Supplementary Table 11]. Scores are indicative of the proportion of DEG and their respective fold changes (negative score – downregulated genes; positive score – upregulated genes) per module

Interestingly, for QIV, the early-stage transcriptome profiles also appeared to associate with antibody persistence profiles (see <u>Materials and Methods</u>) based on the 28-day and 6-month post-vaccination antibody titers [Supplementary Figure 3]. Curiously, for PCV13, the early-stage transcriptome profiles were negatively associated with the antibody persistence profiles [Supplementary Figure 3].

In the case of PCV13, we also stratified individuals by the number of strains against which they mounted a response. The definition of response was adapted from the primary manuscript (*22*) and was based on antibody titers measured 28 days post-vaccination (see <u>Materials and Methods</u>). Based on strain-specific responsiveness, individuals were classified as either Broad responders (responses against more than 11 strains) or Moderate responders (responses against fewer than 9 strains). We observed that individuals with overall higher antibody titers (High responders, quartile 4) also responded to a significantly greater number of strains compared to individuals with overall lower antibody titers (Low responders, quartile 1) [Supplementary Figure 11(a)]. Interestingly, we also noted that the Broad responders showcased high upregulation of inflammation and AP-associated modules compared to Moderate responders at Day 1. However, Moderate responders showed comparable upregulation of inflammation and AP-associated modules at both Day 1 and Day 2.

To investigate transcriptomic differences between overall high and low responders across both vaccines, we re-stratified participants on their dual response type (see <u>Materials and Methods</u>). Briefly, for each vaccine, individuals with quartile scores of 1 or 2 were classified as low responders, whereas those with scores of 3 or 4 were classified as high responders. We then compared post-vaccination transcriptome profiles (PTPs) between individuals who were low responders to both vaccines (overall-low responders) and those who were high responders to both vaccines (overall-high responders) [Supplementary Figure 12]. Overall, the two responder groups exhibited broadly similar activation levels of PTPs across both vaccines. The only notable difference was a significantly greater activation of innate immunity modules in overall-high responders following QIV vaccination.

### Group differences associated with varying levels of major DEGs

The BTM-based approach provides a module-level overview of the immune processes impacted by vaccination. To complement this systems-level perspective, we also examined gene-level expression patterns, focusing on key differentially expressed genes (DEGs) contributing to these immune modules across different comparisons [Figure 6, age, age-corrected frailty groups and antibody responder groups]. To this end, we identified key DEGs on day 1 [Figure 6(a): QIV – Innate response; Figure 6(b): PCV13 – Inflammation response] and day 7 [Figure 6(c): PCV13 – B cell response] post-vaccination that were also reflective of important immune modules (see <u>Materials and Methods</u>). The identified DEGs were linked to specific immune processes via their gene ontology (GO) annotation or literature search [Supplementary Table 8].

**Figure 6:**
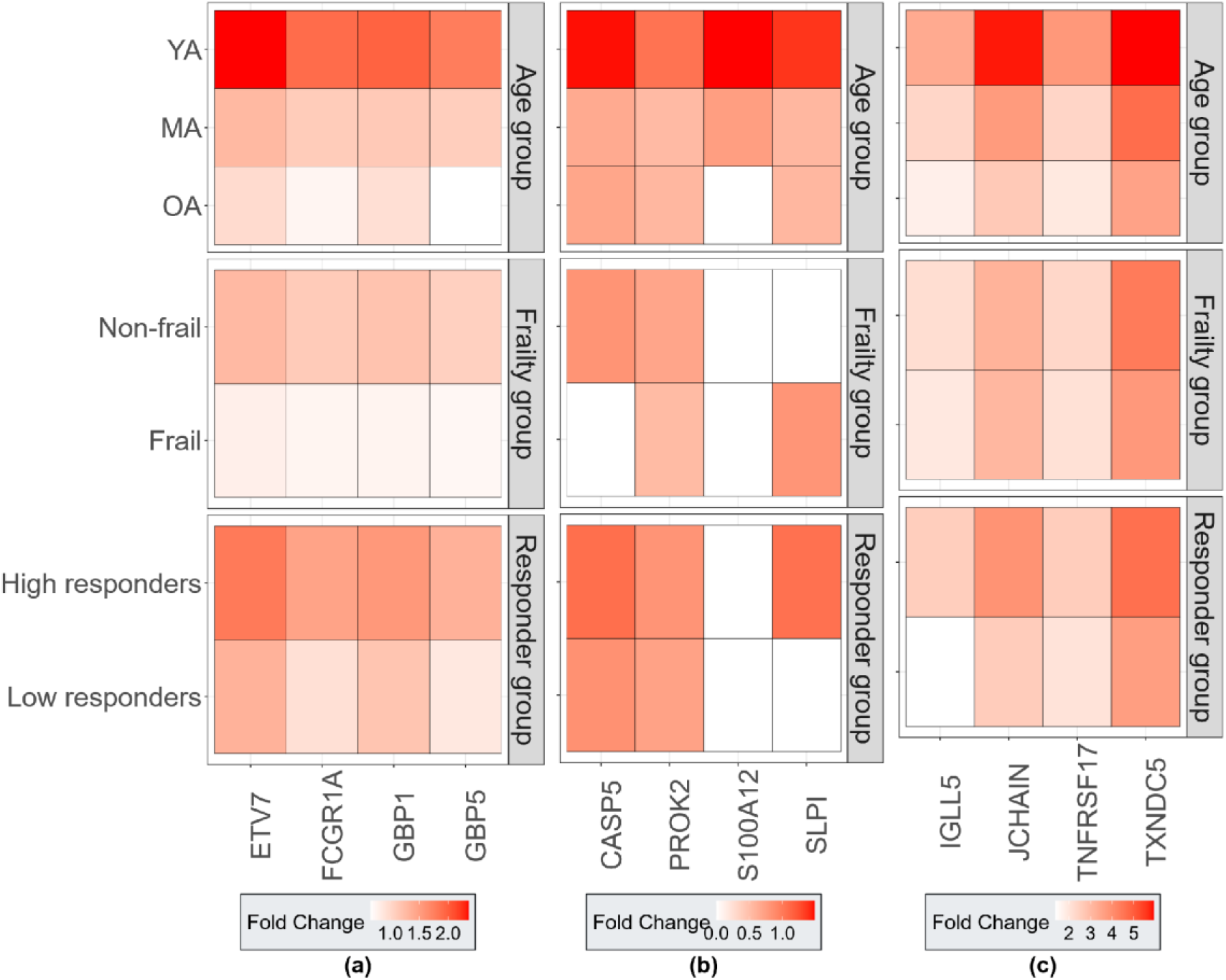
Heatmap plots indicating the differential gene expression for the candidate key DEGs at post-vaccination timepoints compared to the respective pre-vaccination baselines. Panels indicate the genes associated with immune responses at post-vaccination time points, (a) – QIV day-1 Innate immune response, (b) – PCV13 day-1 Inflammation response, (c) – PCV13 day-7 Humoral response. The intensity of colour in each tile is indicative of the magnitude of gene upregulation. In cases where we could not detect a significant differential expression of candidate genes, the tiles are coloured white.

The major differential expression analyses complement the module-based approach by revealing additional signals. For example, in the age-group comparison, both young and older adults showed similar upregulation of the B cell module (*LI.M156.1*: *plasma cells, immunoglobulins*) at day 7 post-PCV13 [Figure 2]. While this reflects shared expression patterns, DEG analysis identified the specific genes (*JCHAIN* and *TXNDC5)* with higher expression in young adults. Overall, both approaches revealed consistent trends: young adults exhibited stronger differential expression of key immune-related genes across both vaccines. Similarly, high antibody responders and non-frail groups showcased higher differential gene expression patterns as compared to low responders and frail groups, respectively. Thus, similar genes contributed to differential gene expression in all groups, but levels of expression were relatively higher in young, non-frail and high antibody response groups.

### Unbiased clustering of the baseline transcriptome aids in identifying groups showcasing strong PTPs

To identify response transcriptome profiles independent of predefined groupings (age, sex, frailty, etc), we applied a less-explored reverse approach (see, (*25*)). Individuals were clustered solely based on similarity in their pre-vaccination (QIV baseline) transcriptome profiles, revealing three distinct baseline transcriptomic clusters (C1, C2, and C3) [Figure 7(a)]. The clustering was driven mainly by expression gradients of specific genes. For instance, at baseline, neutrophil defensin genes were highly expressed in C1 compared to C2 and C3 [Figure 7(a), green box], while interferon-induced genes were elevated in C3 [Figure 7(a), red box]. To further understand the demographic composition of these clusters, we examined the representation of the pre-defined categories within them [Supplementary Figure 2]. We observed an over-representation of OA in C1, MA in C2, and a balanced distribution of OA and MA in C3, with an overall under-representation of YA across all three clusters. Moreover, females were over-represented in C2 and C3, and males in C1.

**Figure 7:**
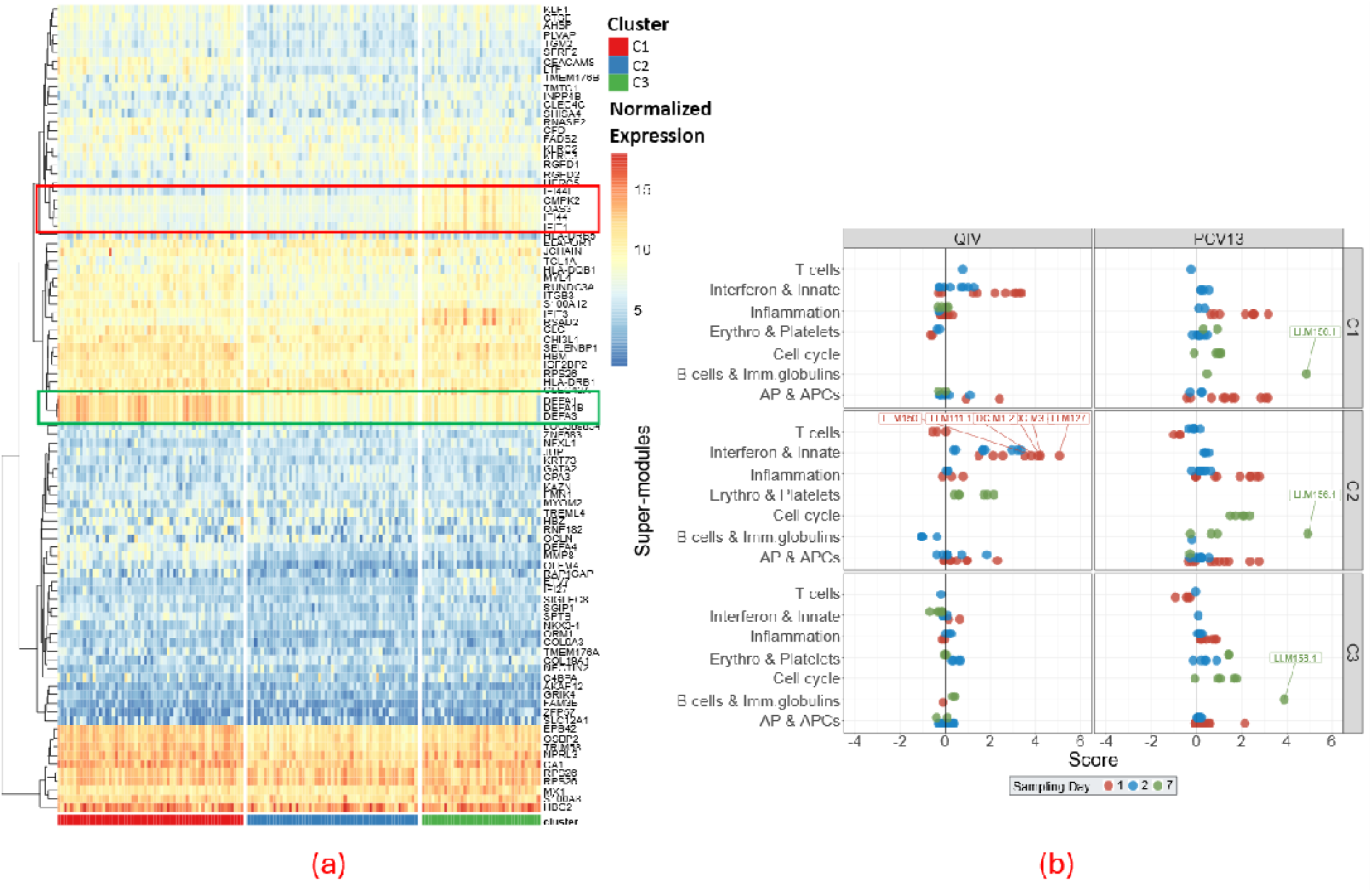
(a) Heatmap depicting the unbiased clusters based on the transcriptome makeup at the flu baseline. (b)Post-vaccination transcriptome profiles (PTPs) for the two vaccines split for the three transcriptome clusters. The impacted BTMs are shown as individual dots and they are scored based on their enrichment of significantly upregulated (positive) or downregulated (negative) genes. The different post-vaccination sampling days are indicated by different colours for the dots. Modules with scores higher than 3.5 are annotated with the Li *et al*., 2014 (*24*) and Chaussabel *et al*., 2008 (*23*) nomenclature [see Supplementary Table 11]. Scores are indicative of the proportion of DEG and their respective fold changes (negative score – downregulated genes; positive score – upregulated genes) per module

Overall, we noted that C3 exhibited relatively weaker PTP than C1 and C2 for both vaccines [Figure 7]. This is further supplemented by the relatively lower DEGs for C3 as compared to C1 and C2 [Supplementary Table 10]. C1 and C2 showed significantly higher upregulation of the interferon, innate response (QIV), and inflammation (PCV13) modules compared to C3 [Supplementary Table 7, BH adjusted p-value < 0.05]. Thus, through unbiased baseline transcriptome categorization, we could identify groups showcasing elevated levels of gene expression in response to both vaccines. The distinct gene signatures elucidated from the baseline transcriptome, which drive the clustering, can be utilized as features to stratify individuals and potentially predict gene expression changes associated with a future vaccine stimulus.

## DISCUSSION

This exploratory study provides new insight into how ageing and frailty shape the early transcriptional response to vaccination. Using a transcriptomics-based approach, we characterized the early gene expression profiles following administration of two vaccines, a booster QIV and primary PCV13, in the deeply phenotyped VITAL vaccination cohort. Both vaccines elicited distinct transcriptional responses involving innate and adaptive immune modules. Differences in PTPs across subgroups (e.g., by age, sex, frailty) were largely attributable to variations in the magnitude of gene expression rather than the expression of distinct gene sets. Notably, both chronological ageing and frailty status, a functional proxy for biological ageing, impacted the vaccine-induced transcriptional changes. Hence, we showed that ageing and frailty exert additive and synergistic influence on the response transcriptome profiles.

QIV induced high upregulation of innate immunity and interferon-based modules, which are consistent with previous findings (*26,27*)[**ICF Nakaya et al 2015**]. Particularly, the induction of the interferon response is in line with an innate response to a viral trigger and serves to prevent viral replication (*28,29*). In contrast, PCV13 induced strong upregulation of early inflammatory pathways, followed by activation of a B cell-associated module (*LI.M156.1*: *plasma cells, immunoglobulins*), which was also in agreement with previous studies (*27,30*). This pattern aligns with the expected immune response to bacterial stimuli, which typically provokes broad inflammatory signalling with a prominent role for macrophages (*31,32*). Despite these differences, both vaccines upregulated antigen presentation-based modules, which are associated with orchestrating immune activation in response to both vaccines and inducing the desired adaptive immune response (*33,34*). Additionally, irrespective of the activated modules, both vaccines showed significant gene expression changes predominantly at day 1, indicating strong immune activation in the first 24 hours post-vaccination. At day 7, PCV13 induced higher upregulation of module *LI.M156.1* (*plasma cells, immunoglobulins*) compared to QIV. This signal may be attributed to the stronger stimulus presented by PCV13, which targets 13 bacterial strains and consequently elicits greater activation of genes linked to B-cell responses compared to QIV. This module is strongly associated with plasmablast expansion and antibody secretion, and has also been highlighted as a universal signature associated with antibody responses at different timepoints (*30*). In our study, we observed that this module was slightly but notably more upregulated in high antibody responders compared to low antibody responders in response to PCV13.

We observed that YA exhibited an overall higher differential expression of immune response modules (QIV – innate immunity and APC-based modules and PCV13 – inflammation-based modules) compared to MA and OA, which was in line with previous studies (*30,35*). The observed decline in transcriptomic responsiveness with age aligns with the reduced efficacy of vaccines in older individuals, with some studies proposing dose optimization to induce robust protection in the older population (see, (*36*)). Ageing is associated with chronic, low-grade inflammation—commonly referred to as *inflammaging*—which is primarily driven by innate immune cells (*37*). This elevated baseline inflammatory state may impair the capacity of the innate immune system to mount a distinct and robust response to vaccination, thereby limiting the transcriptional activation typically seen following immunization. For example, a reduced innate response may result from an age-related decline in the frequency of plasmacytoid dendritic cells (pDCs), a key population of IFN-α–producing cells (*38*). Additionally, only YA showed downregulation of T cell-related modules in response to both vaccines, including module (*LI.M7.4*, *T cell activation(III)*) with genes like *CD96* and *CAMK4*, which are key to T cell activation and expansion (*39,40*). One of the plausible explanations for this downregulation pattern could be the prioritization of innate immune activation over adaptive immune activation. A similar pattern was also observed in a previous study comparing pre-prime to pre-boost timepoints for the inactivated influenza vaccine (*41*). Interestingly, we also note downregulation of CXCR6, a chemokine receptor, which could also be associated with initial T cell activation (*42*).

Importantly, chronological age was not the sole determinant of transcriptomic responses to vaccination, as frailty (as determined with the Frailty Index, (*43*)) also emerged as a significant factor influencing immune responsiveness. Indeed, our previous work demonstrated that frailty has additional effects on inflammaging biomarkers (*11*) and may hamper proper propagation of innate and inflammatory processes. Additionally, older individuals showing evidence of inflammaging will likely have poor immune response to vaccines (*37*). Integrating our findings on age and frailty reveals that even the healthiest older adults, i.e., those with frailty scores comparable to younger individuals, exhibit blunted early transcriptional responses following vaccination as compared to young adults [Supplementary Figure 5]. This pattern suggests that chronological ageing and frailty exert additive and possibly synergistic effects on the immune response, shaping the magnitude of early-stage gene expression changes. Notably, while prior studies have shown that healthy older adults can mount antibody responses similar to those of young adults (*10*), our data highlight that the underlying transcriptional architecture of these responses remains fundamentally altered with age. These findings highlight the need for public health strategies to consider individual health trajectories, not just age, when designing and evaluating vaccination programs for ageing populations.

In addition to age, we also noted significant sex-specific module activation differences in response to QIV exclusively. Specifically, females showed a higher upregulation of the innate immunity-associated modules as compared to males. This observation aligns with previous studies reporting that adult females mount stronger innate responses than males (*44,45*).

Importantly, for QIV, we noted associations between the early-stage transcriptome profiles and the 28-day antibody titers. Specifically, when comparing individuals with low and high antibody titers, we observed that low responders showed lower differential expression patterns compared to high responders, predominantly at day 1. Others have shown that early post-vaccination dynamics in immune cell frequencies, such as plasmablasts and T follicular helper cells, were predictive of antibody responses (*46–48*). In the case of PCV13, we noted a delay in PTPs in low antibody responders, wherein low responders showed similar differential expression of immune response modules as high responders, albeit at day 2. Taken together, for QIV, we observed associations between the early-stage transcriptome profiles and the 28-day and 6-month post-vaccination antibody titers. Such associations were not observed for PCV13. However, further qualitative analyses of PCV13 response revealed that high responders not only mounted an overall high antibody response, but also responded to a broader range of strains than low responders. When stratified by breadth of strain response, we observed that participants responding to more than 11 strains had an overall high upregulation of inflammation and AP-based modules, as compared to participants responding to fewer than 9 strains. Finally, stratification of individuals on their dual response scores revealed comparable levels of PTP activation between the overall-high and overall-low responder groups.

This study also employed an unbiased clustering approach and identified three clusters of individuals based solely on their baseline transcriptome profiles. These clusters exhibited distinct distributions of demographic and clinical variables (age, sex, and responder category) and showed clear differences in post-vaccination transcriptomic responses. The cluster formation was primarily driven by gradients in the expression of specific gene markers predominantly associated with neutrophil defensins and interferon activity [Supplementary Table 6]. Individuals in clusters C1 and C2 demonstrated a generally stronger differential gene expression in response to both vaccines. C3 exhibited elevated expression of interferon-associated genes at baseline and a comparatively attenuated response following vaccination. This association between heightened baseline interferon signatures and diminished vaccine responsiveness has also been reported in previous studies (**ICF Fourati 2022**). Collectively, these findings indicate that baseline transcriptomic signatures, such as elevated interferon-associated gene expression, may reflect intrinsic immune set points that influence vaccine responsiveness, underscoring their potential utility as biomarkers for personalized vaccination strategies in ageing populations.

We used derived variables (antibody response quartiles and age-corrected frailty groups) to quantify transcriptomic differences associated with vaccine response through the lenses of immune response strength (antibody titers) and overall host physiological status (frailty). Adopted from the primary study (*22*), antibody response analyses focused on Quartile 1 (lowest responders) versus Quartile 4 (highest responders), while intermediate quartiles (Quartile 2 and 3) were excluded to maximize contrast between distinct groups. Similarly, age-corrected frailty categories were defined using local regression of the frailty index against age, with individuals near the regression trend excluded to minimize overlap. Without age-correction, differences in transcriptomic profiles would likely reflect age-related biology rather than differences resulting from frailty. Using the regression-based categorisation, we could identify individuals who are “frailer” and “healthier” than expected for their age, thus allowing us to focus on biological variation due to age-independent aspects of frailty. Taken together, focusing on the extreme groups for these derived variables improved the robustness and biological relevance of the comparisons, thereby reducing bias and increasing interpretability.

Our study employed transcriptomic analyses to comprehensively investigate vaccine responses in a uniquely designed ageing cohort, VITAL, in which the same individual was consecutively vaccinated with different vaccines—an approach that distinguishes this work from others (*18–20*). With a robust sample size (n = 148), we were able to stratify participants by demographic and clinical parameters, allowing an in-depth assessment of how these variables influence early gene expression patterns after vaccination. An important limitation of this study was that it did not account for participants’ baseline immunity to the vaccine-targeted pathogens, which would likely be the case for QIV response as participants had previous vaccinations against Influenza. Pre-existing immune status can strongly influence both the magnitude and the quality of post-vaccination responses, and future studies incorporating baseline serological or cellular immunity measures will be essential to comprehensively contextualize transcriptional signatures to obtain a further in-depth understanding of the response patterns. Nevertheless, despite this constraint, our findings reveal vaccine-specific transcriptomic profiles associated with immune responses that advance our understanding of vaccine-induced immunity.

To characterize the post-vaccination expression patterns, we aggregated related BTMs from Li *et al*., 2013 (*24*) and Chaussabel *et al*., 2008 (*23*) into super-modules and quantified their activity using both the proportion and magnitude of differentially expressed genes within each module. Although these sources overlap in terms of genes constituting the modules, neither is a complete subset of the other. From a comparative perspective, this overlap affects all categories similarly and is reflected in the scores. Additionally, the module-based approach relies on group-level differential expression; hence, the resulting scores reflect group-level transcriptional patterns and cannot be extrapolated to the level of individual participants. However, the individual-level variability is accounted for at the gene-level during group-level comparison. Hence, by capturing coordinated activity of related gene sets, the super-module scoring system highlighted broader immunological patterns and revealed vaccine-specific immune signatures. As a supplement, we also compared the expression patterns of genes associated with specific immune processes across the categories [Figure 6]. Additionally, due to logistical reasons, samples at the first post-vaccination time point were not collected on the same day for all participants, resulting in groups sampled either on day 1 or day 2. However, the time points were treated as independent measurements from different experiments rather than interlinked measurements from the same experiment. In addition, the relative proportions of demographic parameters were comparable in the two sampling days [Supplementary Figure 4]. Importantly, for both vaccines, the raw expression levels of the marker genes differed between the two sampling days, with overall higher levels observed on Day 1 [Supplementary Figure 7].

In conclusion, our findings highlight that ageing and frailty status impact the immune response to vaccination. Notably, despite differences induced by the demographic and clinical stratifications, the PTPs shared similar gene signatures across all subgroups. This suggests that the core immune pathways involved in vaccine response remain consistent, but their activation is attenuated in vulnerable populations compared to healthier individuals. These insights point toward future vaccination strategies that could focus more on prioritising to enhance immune responses through dosage optimization or adjuvant additions, rather than identifying group-specific targets, to better support future vaccination strategies tailored for the vulnerable groups.

## MATERIALS AND METHODS

### Study design and participant recruitment

Detailed descriptions of the VITAL study design and recruitment process can be found in the primary manuscript (*22*). A key criterion for recruitment was prior receipt of the seasonal influenza vaccine during the 2018–2019 season. This was done to make the age groups more comparable in terms of their vaccination history. Additional health-related criteria were applied during recruitment to minimize potential confounding factors (*22*). Ethical approval was obtained through the Medical Research Ethics Committee Utrecht (NL69701.041.19, EudraCT: 2019-000836-24). All participants provided written informed consent, and all procedures were performed with Good Clinical Practice and in accordance with the principles of the Declaration of Helsinki. Out of the total 326 participants enrolled in the main cohort, the transcriptomic data were generated for 148 individuals.

Due to logistical constraints, participants were divided into two groups for the first post-vaccination collection (day 1 or day 2). Although the number of individuals sampled differed between these two days (day 1: n=86; day 2: n=62), the relative proportions of sexes and age categories were comparable [Supplementary Figure 4]. These day-specific groupings for the first post-vaccination timepoint were consistent across both vaccines and are taken into account in the analyses.

Selection of individuals within the transcriptome sub-cohort was decided based on the following criteria:

- Informed consent from participants for in-depth transcriptome (DNA/RNA) analysis
- Availability of antibody data against both vaccines
- Availability of whole blood stored in paxgene tubes at pre-vaccination and post-vaccination (Day 1/2 and 7) timepoints for both vaccines
- Post-vaccination Day 1 or Day 2 alignment for both vaccines

These criteria were defined post-hoc. Finally, due to the unavailability of data on either of the two vaccines for two participants, the transcriptome analyses focused on 148 individuals. The study setup is outlined in [Figure 8]

**Figure 8:**
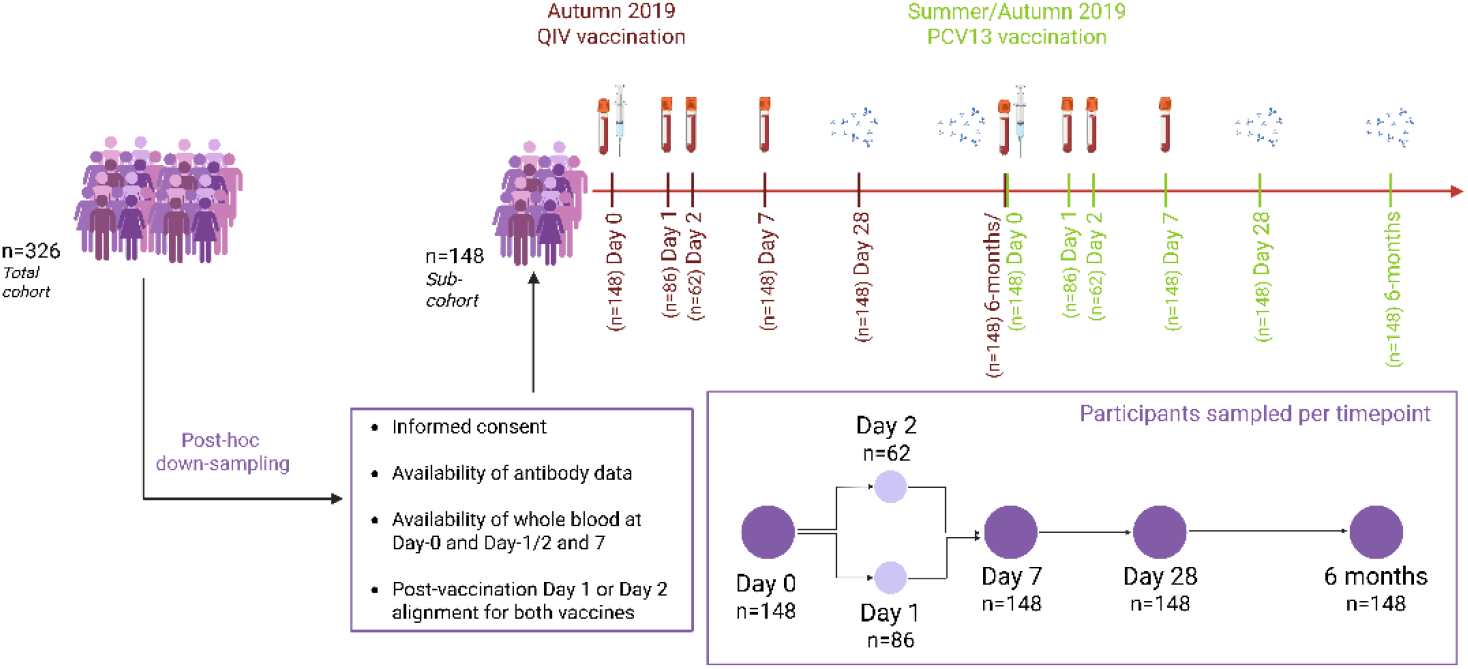
Study setup of the transcriptome sub-cohort. Of the total 326 participants, 148 were shortlisted based on post-hoc down-sampling criteria. Transcriptome dataset was generated for whole blood samples collected at pre-vaccination baseline (Day-0) and post-vaccination (Day-1, Day-2 and Day-7) timepoints for both vaccines. Additionally, antibody data collected at Day-28 and 6-month timepoints were used for identifying associative patterns between antibody response and transcriptome profiles.

### Vaccinations and sample collections

During the autumn of 2019, all participants were initially vaccinated with the seasonal quadrivalent influenza vaccine (QIV, Influvac Tetra, Abbott Biologicals B.V. The Netherlands), which contained neuraminidase and hemagglutinin from the following viral strains: A/Brisbane/02/2018, IVR-190(H1N1); A/Kansas/14/2017, NYMC X-327 (H3N2); B/Maryland/15/2016, NYMC BX-69A (B/Victoria/2/87 lineage); and B/Phuket/3073, wildtype (B/Yamagata/16/88 lineage). Following this, in summer/autumn 2020, all participants received the 13-valent pneumococcal polysaccharide conjugate vaccine (PCV13, Prevnar 13, Pfizer Europe, Belgium) containing polysaccharides from serotypes 1, 3, 4, 5, 6A, 6B, 7F, 9V, 14, 18C, 19A, 19F and 23F conjugated to CRM197 carrier protein.

Whole blood samples were collected at six timepoints: Flu – day 0 (pre-vaccination), day 1 or 2 and day 7 (post-vaccination); PCV13 – day 0 (pre-vaccination), day 1 or 2, and day 7 (post-vaccination).

### Vaccine-specific antibody titers and stratification

Vaccine-specific antibody response scores were adopted from the primary study (*22*). Briefly, antibody levels measured at 28 days post-vaccination (QIV: H3N2 titers; PCV13: IgG concentrations against all 13 serotypes) were divided into quartiles. Since H1N1 titers were high at baseline and did not reveal strong responder vs non-responder distinctions, only H3N2 titers were used for QIV stratification. Individuals were then assigned a score from 1 to 4 based on their antibody response, with 1 representing the lowest quartile (low responders), 4 the highest quartile (high responders), and 2–3 the intermediate quartiles (intermediate responders). In the case of PCV13, quartile scores were determined separately for each of the 13 serotypes, and the commonly occurring quartile score was assigned as the individual’s overall quartile score. To maximize the contrast, we focused on studying PTPs of low and high responders only.

Using the vaccine-specific quartile scores for the two vaccines, we next assigned a dual response type to each participant. Individuals with low quartile scores (1–2) for both vaccines were classified as overall-low responders, whereas those with high quartile scores (3–4) for both vaccines were classified as overall-high responders. Participants exhibiting discordant responses between the two vaccines (i.e., low for one vaccine and high for the other; n = 80) were excluded from this analysis.

For PCV13-based qualitative analysis, individuals were categorized based on the number of PCV13 strains to which they responded: Moderate responders (<9 strains) and Broad responders (>11 strains). Here, the definition of response was adapted from the primary study (*22*): antibody titer ≥1.3 µg/mL at 28 days post-PCV13 vaccination together with at least a twofold increase relative to the pre-vaccination baseline.

To quantify antibody response durability, a persistence score was calculated for each individual using antibody measurements obtained at baseline, 28 days, and 6 months post-vaccination [equation 1]

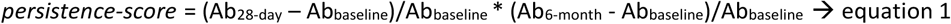

Similar to the quartile-based classification of antibody titers, individuals were assigned a quartile score based on their persistence score per vaccine. PTPs were then compared between individuals with the highest (high-responders) and lowest (low-responders) persistence scores.

### Transcriptomics data generation and quality control

The transcriptomic dataset was generated from whole blood samples collected in Paxgene tubes (Qiagen). Before RNA extraction, samples were randomized by age group. RNA was extracted from 897 samples using the Pre-Analytix PAXgene Blood RNA Kit. The samples originating from pre- and post-vaccination timepoints for the two vaccines were normalized together to avoid bias. Total RNA concentration and quality were assessed using a NanoDrop ND-8000 spectrophotometer and an Agilent 2100 Bioanalyzer, respectively. Transcriptomic profiling of all samples was performed by Charles River Laboratories using the Affymetrix Clariom™ S Array HT platform. Arrays were scanned using the GeneTitan Instrument, and raw data were processed with the Affymetrix GeneChip® Command Console (AGCC).

Quality control (QC) was conducted using Transcriptome Analysis Console (TAC) software (version 4.0), with data summarized using SST-RMA (Signal Space Transformation–Robust Multi-array Average). QC parameters (total of 49) were monitored for the extraction step (RNA quality (n=10 RIN <4) and quantity (n=12 concentration < 33ng/µl), the labelling & hybridization step (Gene titan platform, cRNA yield (n=1 < 15 µg), ssDNA yield (n=0 < 5.5 µg)), and for Affymetrix Transcriptome Analysis Platform QC step (back-ground correction, normalization, summarization, QC thresholds: Image issues (n=51), Control hybridation (n=0), Low Signal BoxPlots (n=6), positive vs negative probes area under the curve (AUC) > 0.7 (n=0), all probeset RLE mean > 0.6 (n=26)). Overall, 153 QC flags were raised (and 97 excluding minor comments) for 897*49 QC parameters measured. The impact of these flags on data quality was further assessed using an intensity-based multivariate analysis with the Array Quality Metrics QC package (ICF Kauffmann 2009) on three criteria that raised 54, 5, and 0 flags, respectively. Considering a sample exclusion, as done in the Human Immunology Project Consortium (HIPC; (*51*)), zero samples were to be removed.

We further investigated three other common outlier detection methods, based on PCA (P99 distance), *lumi* package method (*52*), and the Correlation-based QC based Median Absolute Deviation scores, which raised respectively 9, 27 and 45 QC flags. Based on these results, we removed nine samples based on the primary criteria to keep baseline samples. The generated transcriptome dataset is deposited in the *ArrayExpress* database under the following accession identifier: E-MTAB-16338.

### Bioinformatics and statistical analyses

The majority of the bioinformatics analyses were performed using the *R* programming language ((*53*), version 4.4.3). Batch effects were corrected with *ComBat* function from the *sva* package (version 3.50.0) with parametric empirical Bayesian adjustments. Differential gene expression analysis was performed using a mixed Gaussian model for repeated measures on log2-transformed expression values. The model included subject as a random factor, and time, response, and their interaction as fixed effects, implemented via the *dream* function from the *variancePartition* package (version 1.32.5). Adjustments for multiple testing were made using the false discovery rate (FDR) approach of *Benjamini and Hochberg*(*54*) at a nominal level of 5%. Gene annotation was performed using the *clariomshumantranscriptcluster.db* package (database version 8.8.0).

Differentially expressed genes (DEGs) were identified using a p-value threshold of 0.05, without applying a log-fold change cutoff. All DEGs were derived by comparing gene expression profiles at post-vaccination timepoints with pre-vaccination (baseline) levels. These DEGs stem from contrasting demographic, clinical and transcriptome-variables at different timepoints. To exemplify, (D7 – D0)_High responders_ indicate the DEGs at Day 7 compared to Day 0 (baseline) for high vaccine responders. The two vaccines had independent baselines [<u>see</u> Figure 8]. Majorly differentially expressed genes (DEGs) were associated with specific immune responses through gene ontology terms or literature searches [Supplementary Table 8].

On the level of functional pathways, we used the blood transcriptome module (BTM)-based gene categorization to characterize the post-vaccination transcriptome profiles (PTPs). Specifically, we made use of the gene categories as described in Li *et al*., 2014(*24*) and Chaussabel *et al*., 2008(*23*) that were derived using the *tmod* package (version 0.50.13, (*55*)).

### Grouping and scoring transcriptome modules into *super-modules*

The BTMs used in this study were derived from Li *et al*., 2013 (*24*) and Chaussabel *et al*., 2008 (*23*). To obtain a systems-level view of immune changes post-vaccination, related BTMs were grouped into eight *super-modules*. Here, super-modules are catalogues of similar BTMs that are all reflective of specific immune processes. Hence, super-modules capture cohort-level summaries of coordinated transcriptional changes. Therefore, these catalogues can be used to obtain an overview of the vaccine responses based on enrichment patterns of DEGs within them. The constructed super-modules and the modules contained within them are listed in [Supplementary Table 9]. Individual modules were scored based on the proportion of differentially expressed genes (DEGs) and their respective fold changes using equation 2:

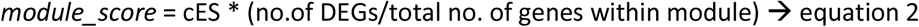

Here, *cES* is indicative of the cerno statistic extracted from *tmod* package. In short, a higher module score would indicate a strong enrichment of differentially expressed genes within a given BTM. To incorporate directionality, genes within the modules were split based on their direction of differential expression and scored separately. Modules enriched for upregulated genes were given a positive score, whereas those enriched for downregulated genes were given a negative score. Hence, a positive module score indicates significant upregulation of genes associated with the module, with the magnitude reflecting the strength of enrichment and the proportion of differentially expressed genes contributing to the module signal.

Admittedly, the modules are not independent due to shared genes and coordinated biological functions. Nevertheless, to obtain an overall view of module activity across comparisons, we performed pairwise Welch’s t-tests to assess the distributions of module scores.

### Age-corrected frailty analysis

To ensure the frailty index was applied in a clinically and biologically meaningful context, we limited our analysis to individuals aged 50 years and older. Frailty in younger adults is rare and less predictive of adverse health outcomes, as supported by the original validation studies of the frailty index by Mitnitski *et al.,* 2001 (*43*). Therefore, including only participants aged 50 or above makes comparisons between frailty groups more robust and interpretable. Using the precalculated individual-specific frailty indices (*11,22*), we performed local regression analysis to model the relationship between chronological age and frailty index (see Figure 4a). The frailty score was based on the Rockwood and Mitnitski methodology (*56*). The resulting regression curve served as an age-specific predictor for frailty, allowing us to classify individuals into two groups: age-corrected frail (frailty index above the regression curve) and age-corrected non-frail (frailty index below the curve). Individuals with frailty indices falling within the confidence interval of the regression line were excluded from this age-corrected classification.

### Baseline transcriptome clustering

Using exclusively the baseline (pre-vaccination QIV, T_0_) transcriptome dataset, we cluster individuals to link baseline gene expression patterns with post-vaccination response patterns. Specifically, we first identified the top 100 genes from baseline with the highest standard deviation in their absolute gene expression values using the *probe_ranking* function from the *multiClust* package (version 1.32.0). We then used the *omada* package (version 1.4.0) to cluster individuals based on the expression values of these selected genes. Internally, the package applies three clustering techniques—hierarchical, spectral, and k-means. Based on the highest partition agreement score, we selected *n = 3*. By using distinct timepoints for clustering and analyses, we avoid information leakage (statistical double-dipping)

## Author contributions

Conceptualization: D.v.B, J.v.B, D.L., W.B., N.R., & A.B.

Methodology and Approach: C.C., E.C., V.C., M.C.R., L.B., & M.J.

Data management: E.B.

Investigation: M.J. & C.C.

Visualization and interpretation: M.J., C.C., M.v.H., A.C., Y.v.S, D.v.B., D.L., W.B., & M.C.R

Writing: M.J., Y.v.S., M.v.H., D.v.B., C.C., M.C.R., & V.C.

Review: All authors

## Data Availability

The analyses were carried out using an in-house developed pipeline designed using the R programming language. This pipeline is stored in the following GitHub repository: https://github.com/Manaswwm/TRAVIT. The transcriptome dataset is hosted as a private ArrayExpress dataset (accession ID - E-MTAB-16338), access will be made public upon acceptance of the manuscript in a peer-reviewed journal.

## List of abbreviations

QIV: Quadrivalent Influenza Vaccine
PCV13: 13-valent Pneumococcal Conjugate Vaccine
VITAL: **V**accines and **I**nfec**T**ious diseases in the **A**geing population
BTMs: Blood Transcriptome Modules
PTPs: Post-vaccination Transcriptome Profiles
YA: Young Adults
MA: Middle-aged Adults
OA: Older adults
DEGs: Differentially Expressed Genes
AP & APCs: Antigen Presentation and Antigen Presenting Cells
GO: Gene Ontology
C1, C2, C3: Cluster1, Cluster2, Cluster3
TC: Transcriptome cohort
Non-TC: Whole cohort Transcriptome Cohort

## SUPPLEMENTARY MATERIAL

**Supplementary Figure 1:**
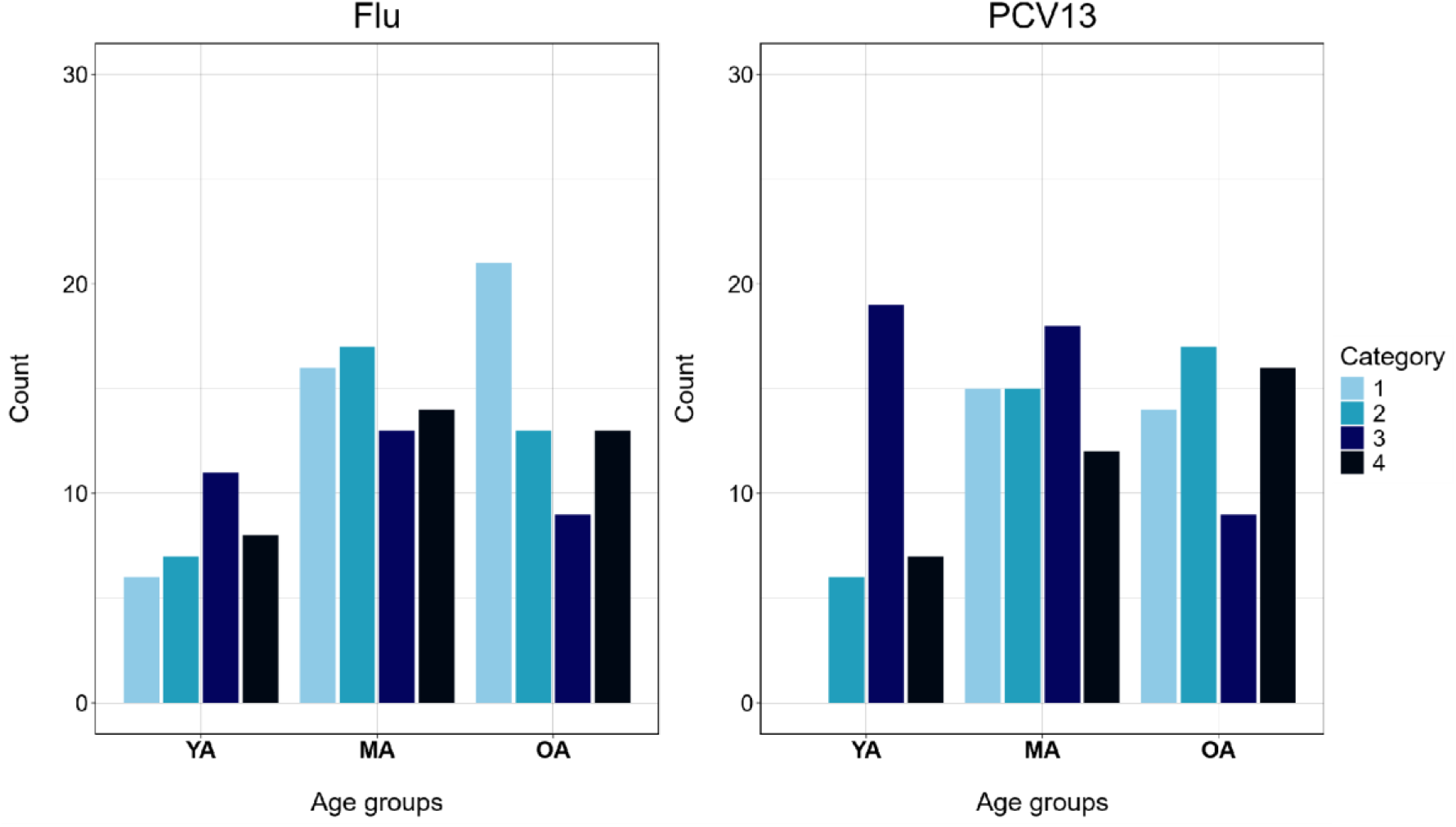
Distribution of age categories per responder stratification groups for the two vaccines.

**Supplementary Figure 2:**
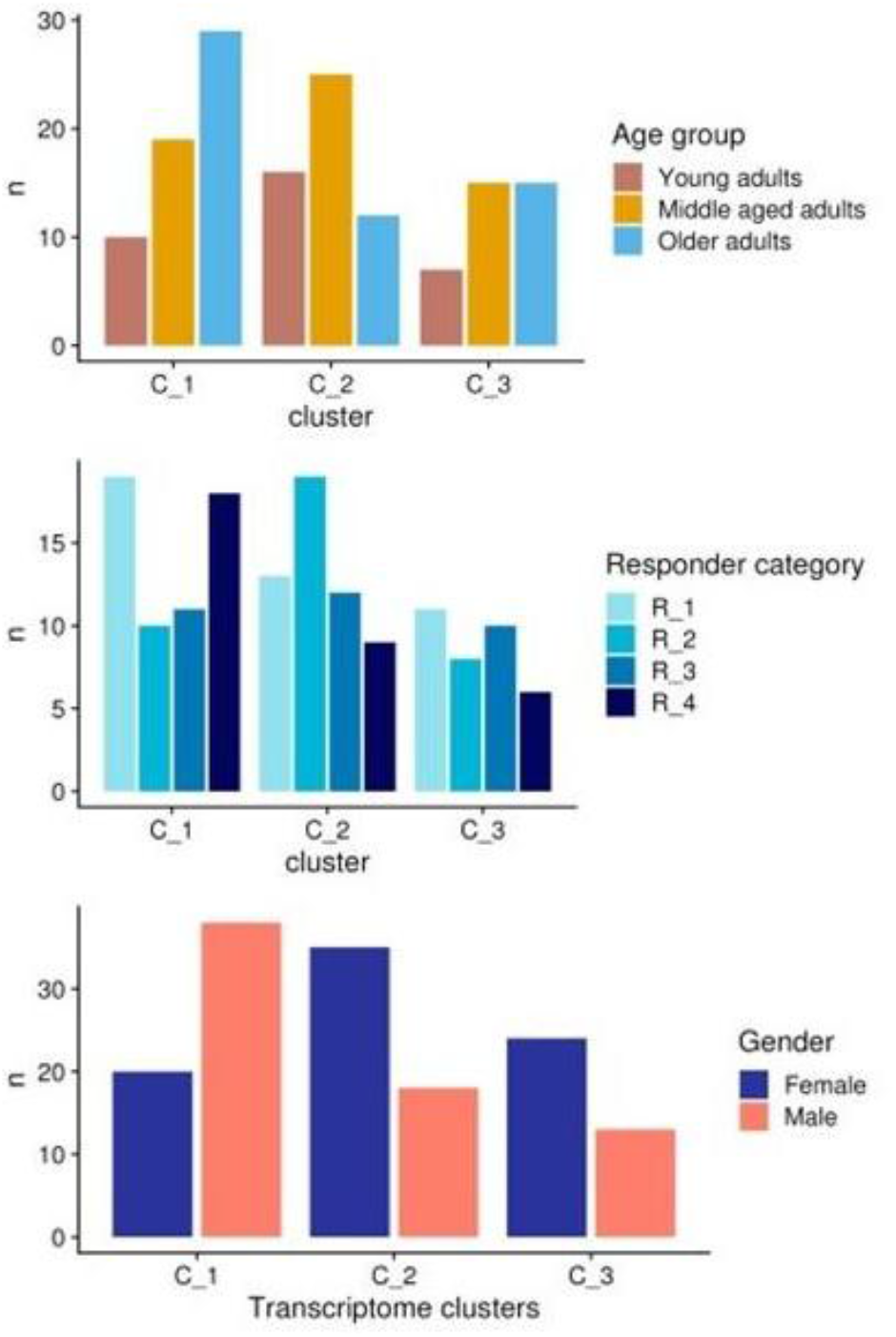
Distribution of demographic and clinical parameters (age, sex and response stratification) of the three unbiased transcriptome clusters

**Supplementary Figure 3:**
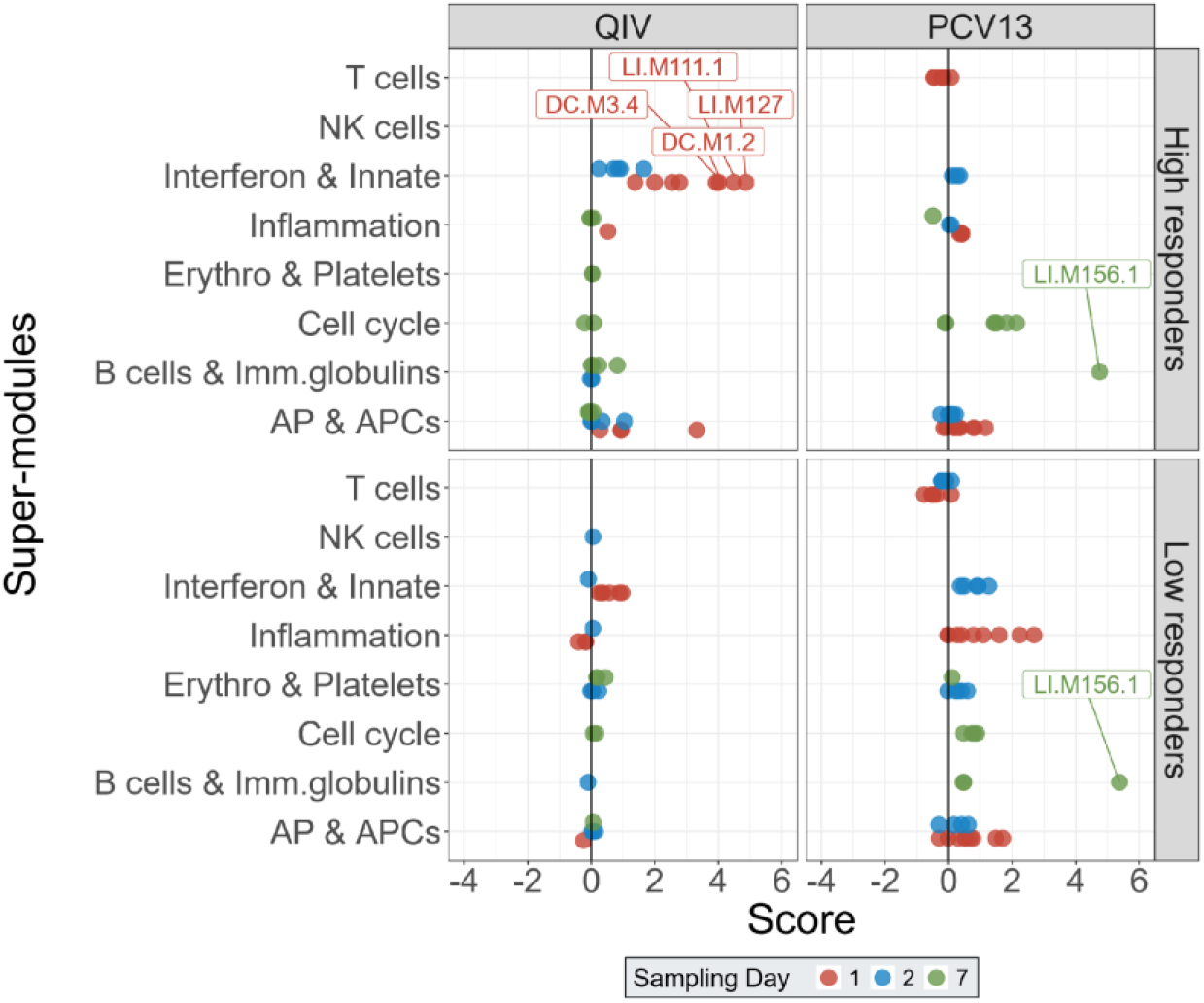
BTM-based analyses of the two antibody persistence groups (high and low responders) for both vaccines based on the 28-day and 6 month post-vaccination titers. Persistence scores are derived using equation 1. In brief, a higher score indicates relatively higher titers at both timepoints combined, while lower score indicates lower titers. Score (X-axis) is indicative of the levels and direction (up/down) of the regulation of modules. Super-modules (Y-axis) are collection of individual BTMs that participate in a specific immune process. The colors of the datapoints are indicative of the sampling days post-vaccination. Modules with scores higher than 3.5 are annotated with the Li *et al*., 2014 (*24*) and Chaussabel *et al*., 2008 (*23*) nomenclature [see Supplementary Table 12]

**Supplementary Figure 4:**
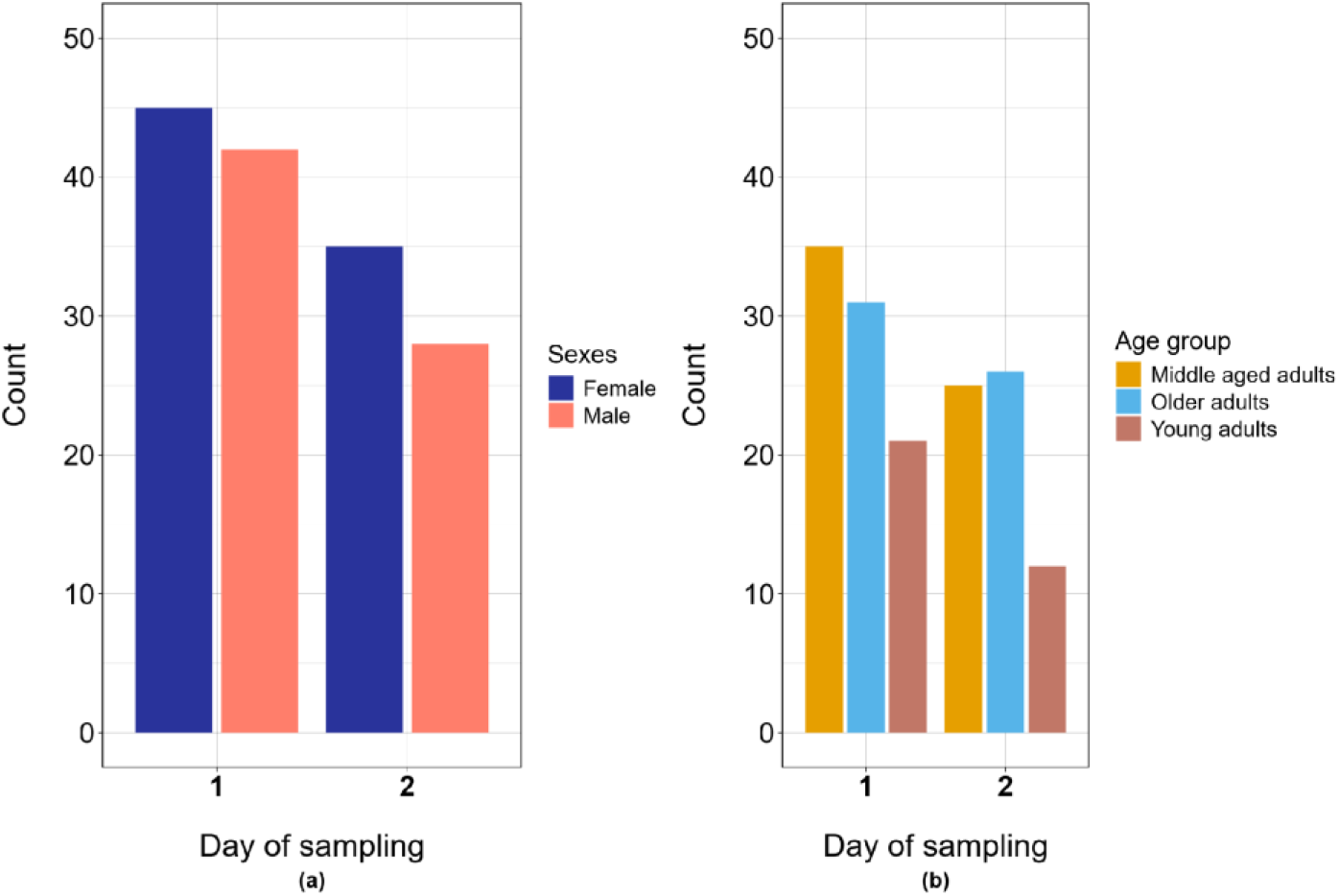
Distribution of the sexes (a) and the age groups(b) at the two sampling days for the first timepoint post-vaccination. The distributions were consistent for both vaccines.

**Supplementary Figure 5:**
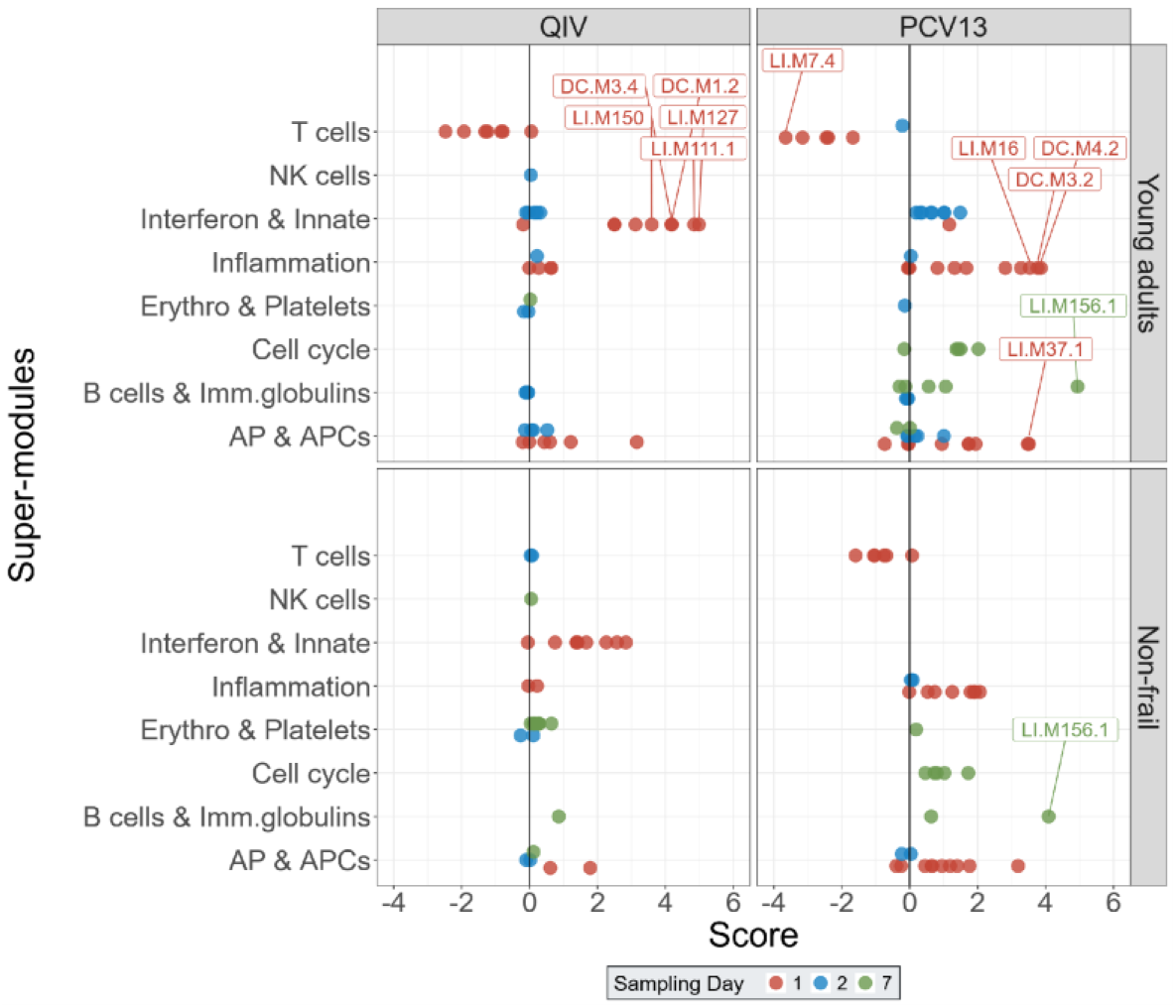
The PTPs for the two vaccines split for Young adults and Non-frail groups. The individual BTMs are indicated as a single dot and are coloured based on the sampling timepoint post-vaccination. The modules are scored based on the enrichment of genes significantly upregulated (positive score) or downregulated (negative score). Modules with scores higher than 3.5 are annotated with the Li *et al*., 2014 (*24*) and Chaussabel *et al*., 2008 (*23*) nomenclature [see Supplementary Table 12]

**Supplementary Figure 6:**
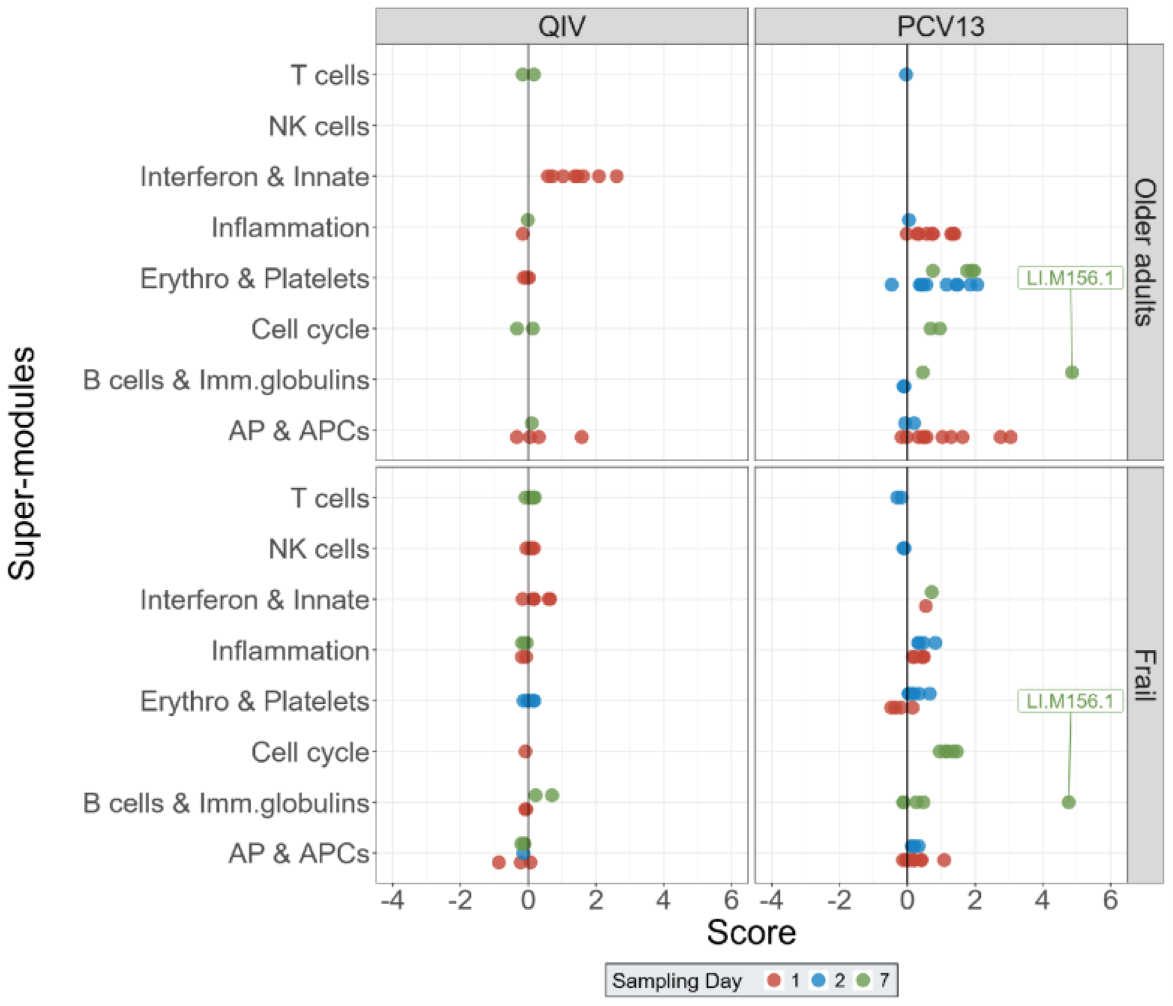
The PTPs for the two vaccines split for Older adults and Frail groups. The individual BTMs are indicated as a single dot and are coloured based on the sampling timepoint post-vaccination. The modules are scored based on the enrichment of genes significantly upregulated (positive score) or downregulated (negative score). Modules with scores higher than 3.5 are annotated with the Li *et al*., 2014 (*24*) and Chaussabel *et al*., 2008 (*23*) nomenclature [see Supplementary Table 12]

**Supplementary Figure 7:**
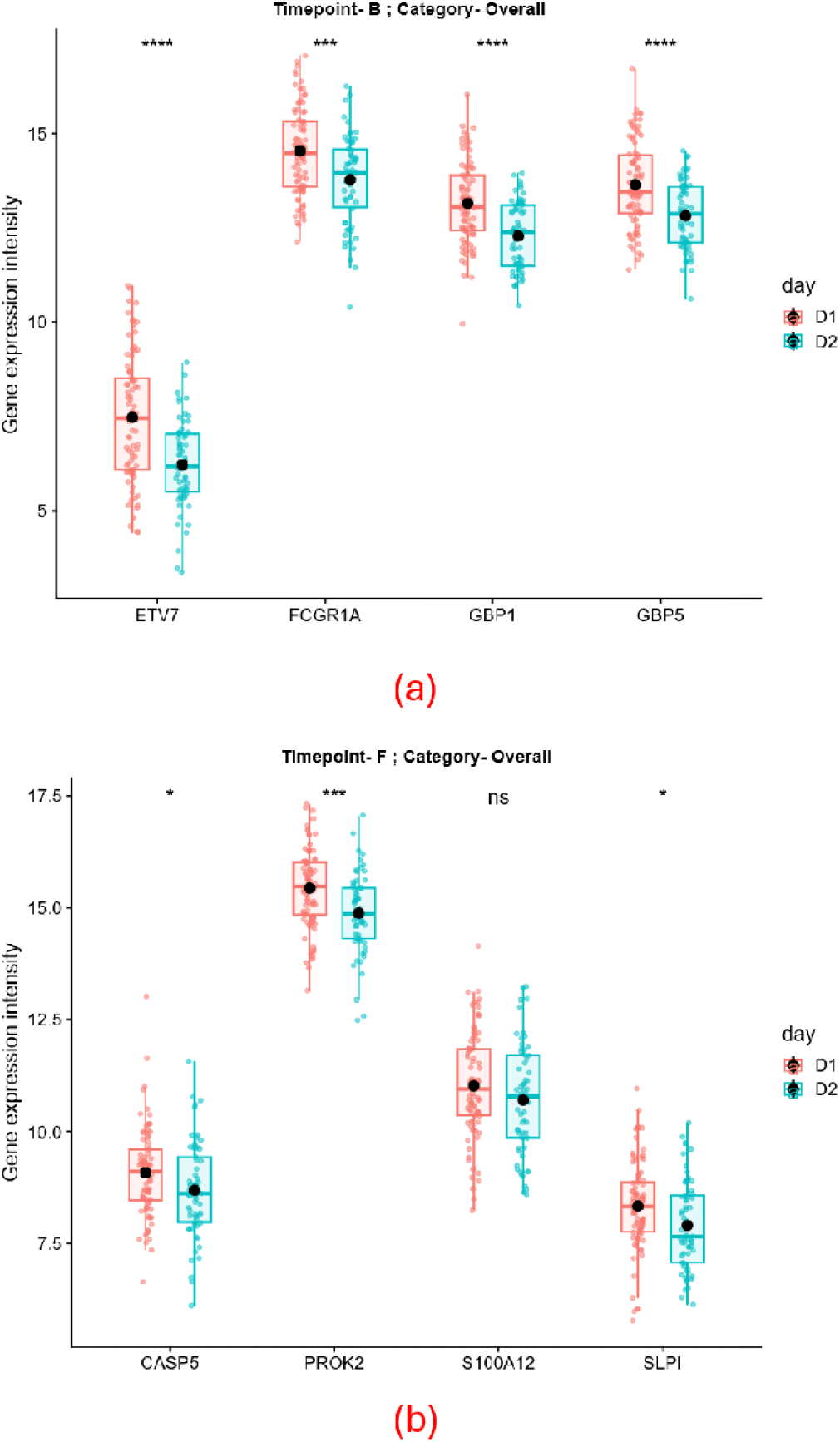
Comparing the raw gene expression values of the marker genes for flu (a, Timepoint-B) and PCV13 (b, Timepoint-F) at sampling days 1 and 2 using t-test. The p-value significance are indicated with symbols (ns – non-significant, * - p-value < 0.05, ** - p-value <0.01, *** - p-value < 0.001, **** - p-value < 0.0001), mean values are indicated with black circle

**Supplementary Figure 8:**
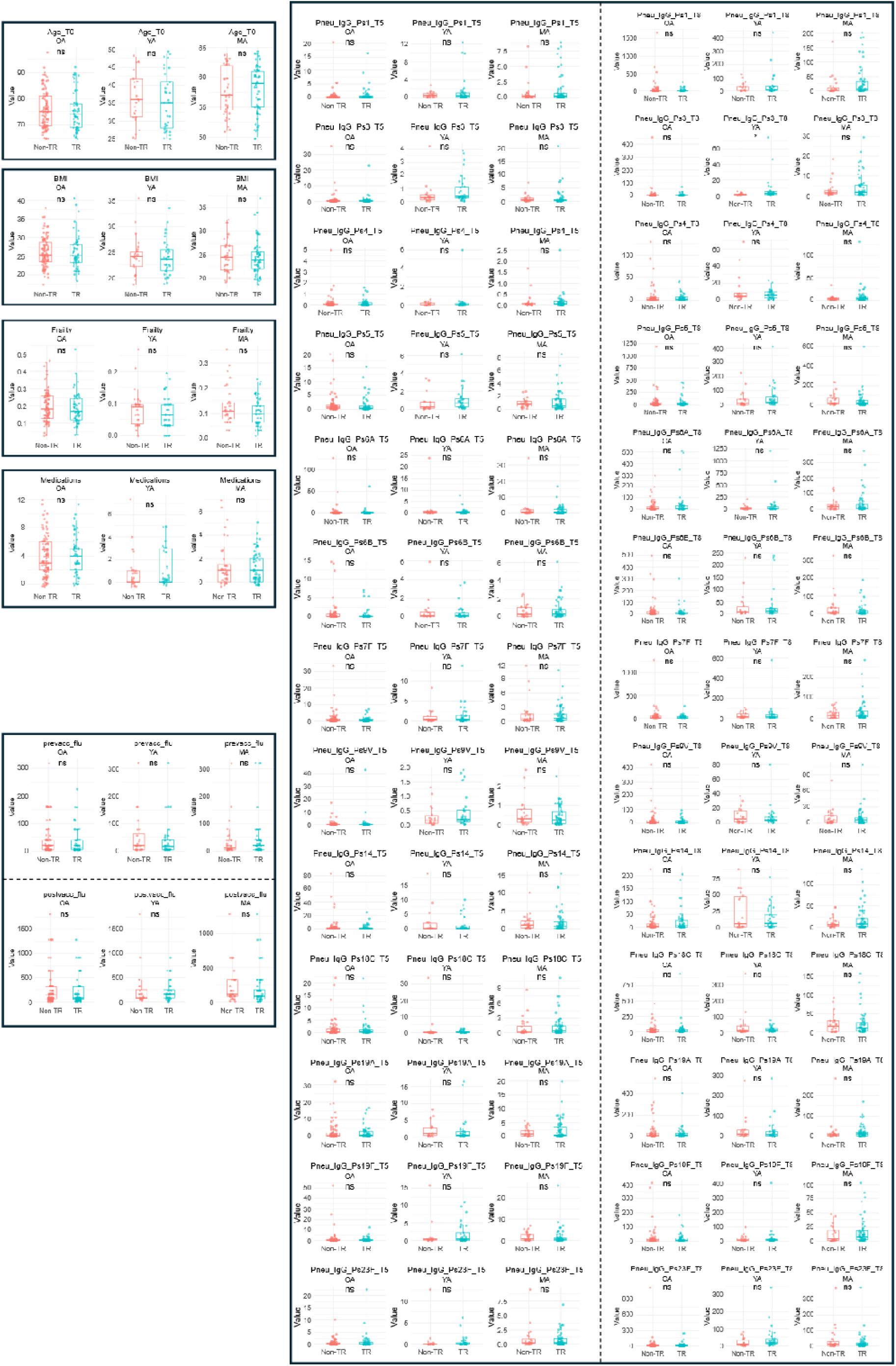
Demographic and clinical parameter comparisons in the main (Non-TR) and the transcriptome sub-cohort (TR). Here, Non-TR excludes the participants within the TR sub-cohort. Parameters – age (Age_T0), Frailty index (Frailty), BMI (BMI), no. of medications (Medications), QIV baseline (prevacc_flu), QIV post-vaccination (postvacc_flu), PCV13-baseline (Pneu_IgG_*_T5) and PCV13 post-vaccination (Pneu_IgG_*_T8). Significance values are derived post correction (method – Benjamini Hochberg)

**Supplementary Figure 9:**
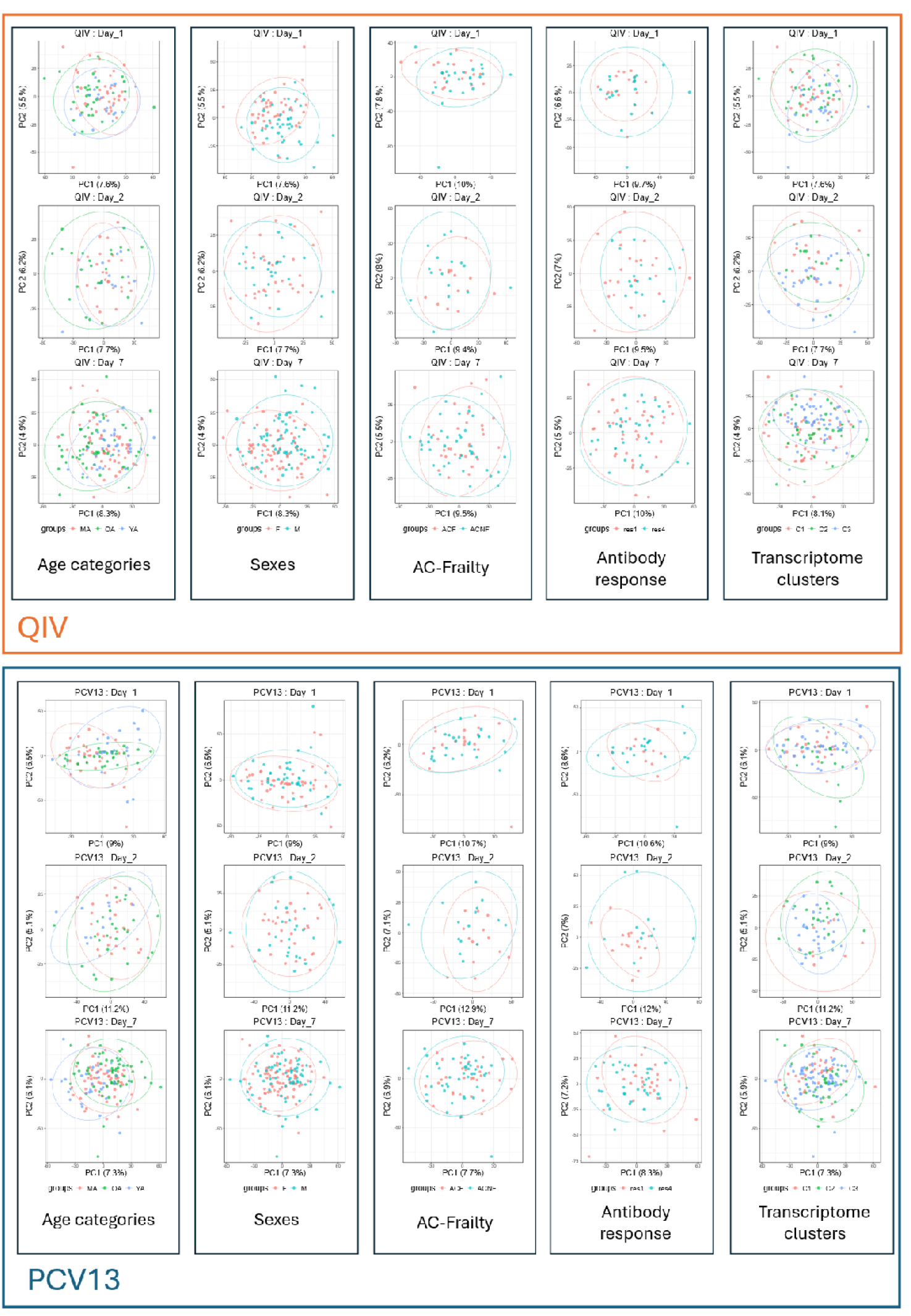
Principal component analysis (PCA) of post-vaccination timepoints compared to pre-vaccination baseline across stratifications for the two vaccines. Stratification guide: MA (middle-aged adults), OA (older adults), YA (young adults), F (Females), M (Males), ACF (age-corrected frail), ACNF (age-corrected non-frail), res1 (low responders), res4 (high responders), C1 (transcriptome cluster 1), C2 (transcriptome cluster 2), C3 (transcriptome cluster 3)

**Supplementary Figure 10:**
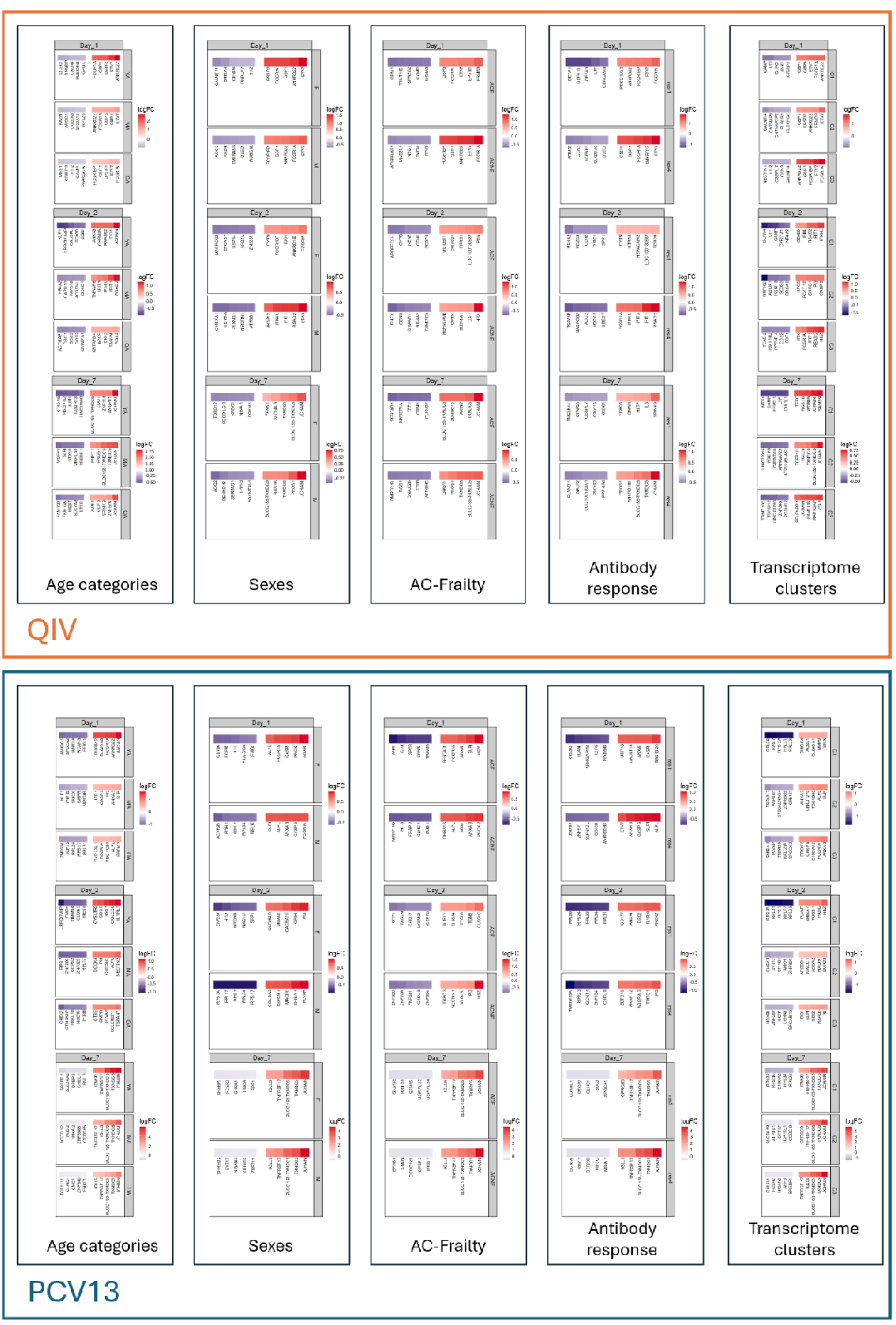
Top 5 up- and down-regulated genes of post-vaccination timepoints compared to pre-vaccination baseline across stratifications for the two vaccines. Stratification guide: MA (middle-aged adults), OA (older adults), YA (young adults), F (Females), M (Males), ACF (age-corrected frail), ACNF (age-corrected non-frail), res1 (low responders), res4 (high responders), C1 (transcriptome cluster 1), C2 (transcriptome cluster 2), C3 (transcriptome cluster 3)

**Supplementary Figure 11:**
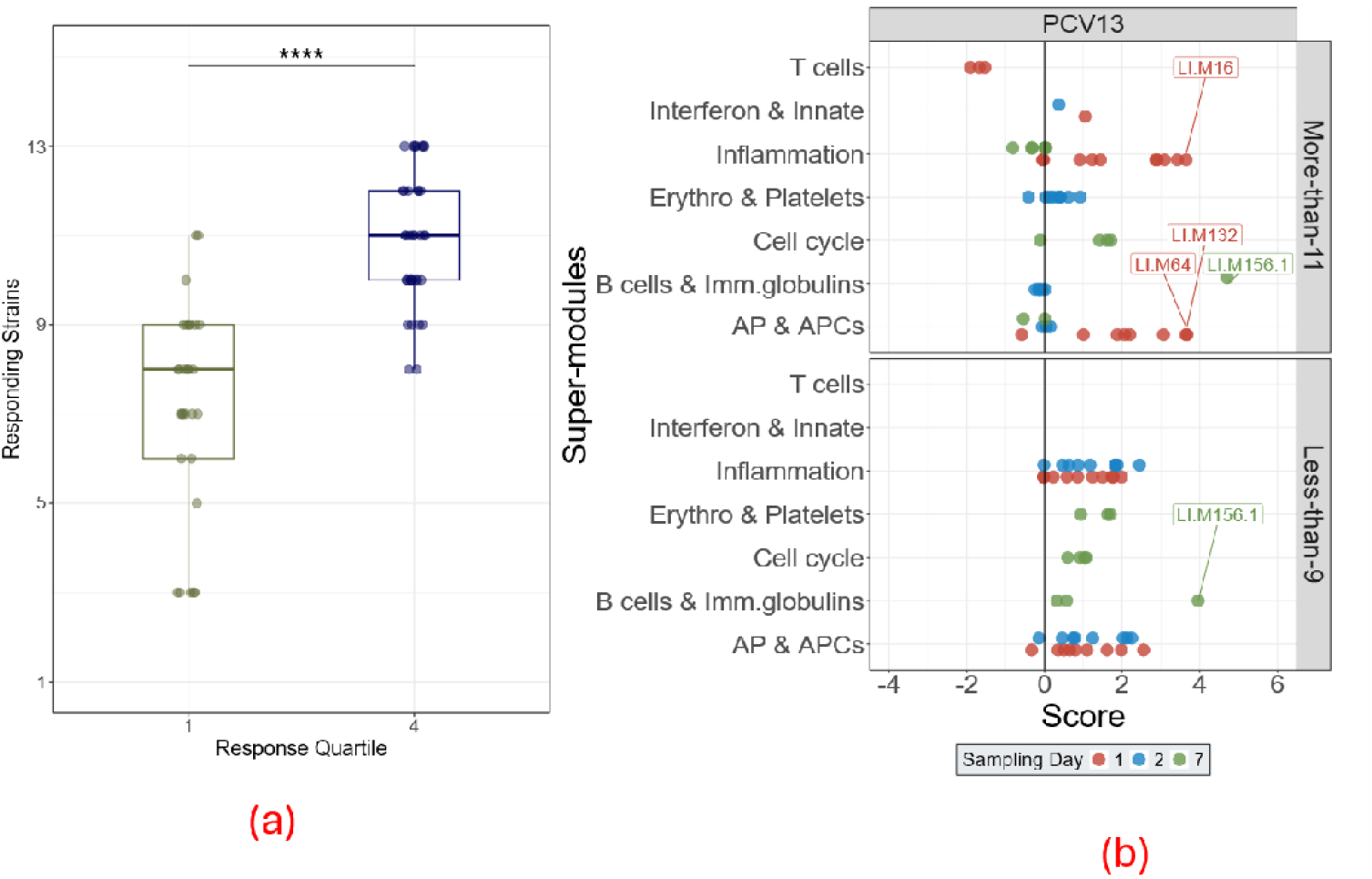
Stratifying individuals based on the number of PCV13 strains individuals responded to, response criteria is described in the primary manuscript (22). In short, response is defined as antibody titer ≥1.3 µg/mL at 28 days post-PCV13 vaccination together with at least a twofold increase relative to the pre-vaccination baseline. (a) – Number of strains individuals responded to stratified by the response quartiles. Significance of difference in distribution is derived on adjusted p-value (b) – BTM-based analysis of individuals stratified on the number of strains individuals responded to: group 1 (more than 11 strains) and group 2 (less than 9 strains)

**Supplementary Figure 12:**
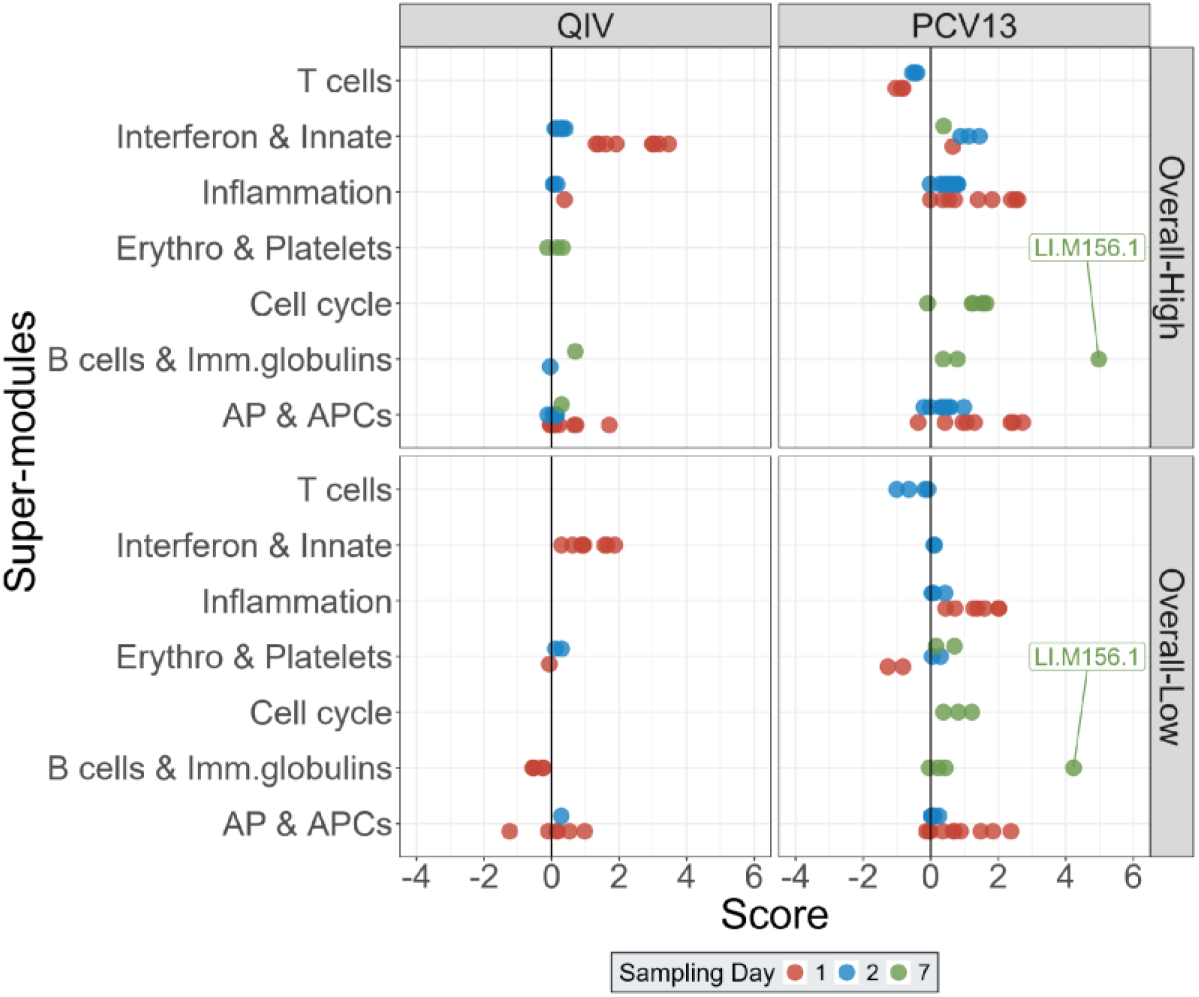
BTM-based analysis of individuals stratified based on the combined vaccine response to the two vaccines. In short, based on 28-day post-vaccination antibody titers, individuals having quartile score of 1 or 2 were defined as overall low responders (Overall-Low), while those having quartile score of 3 or 4 were defined as overall high responders (Overall-High). This category assignment was done per vaccine. Individuals who were either Overall-High or Overall-Low for both vaccines were used in the analysis (n=68), the rest were omitted (n=80).

**Supplementary Table 1:**
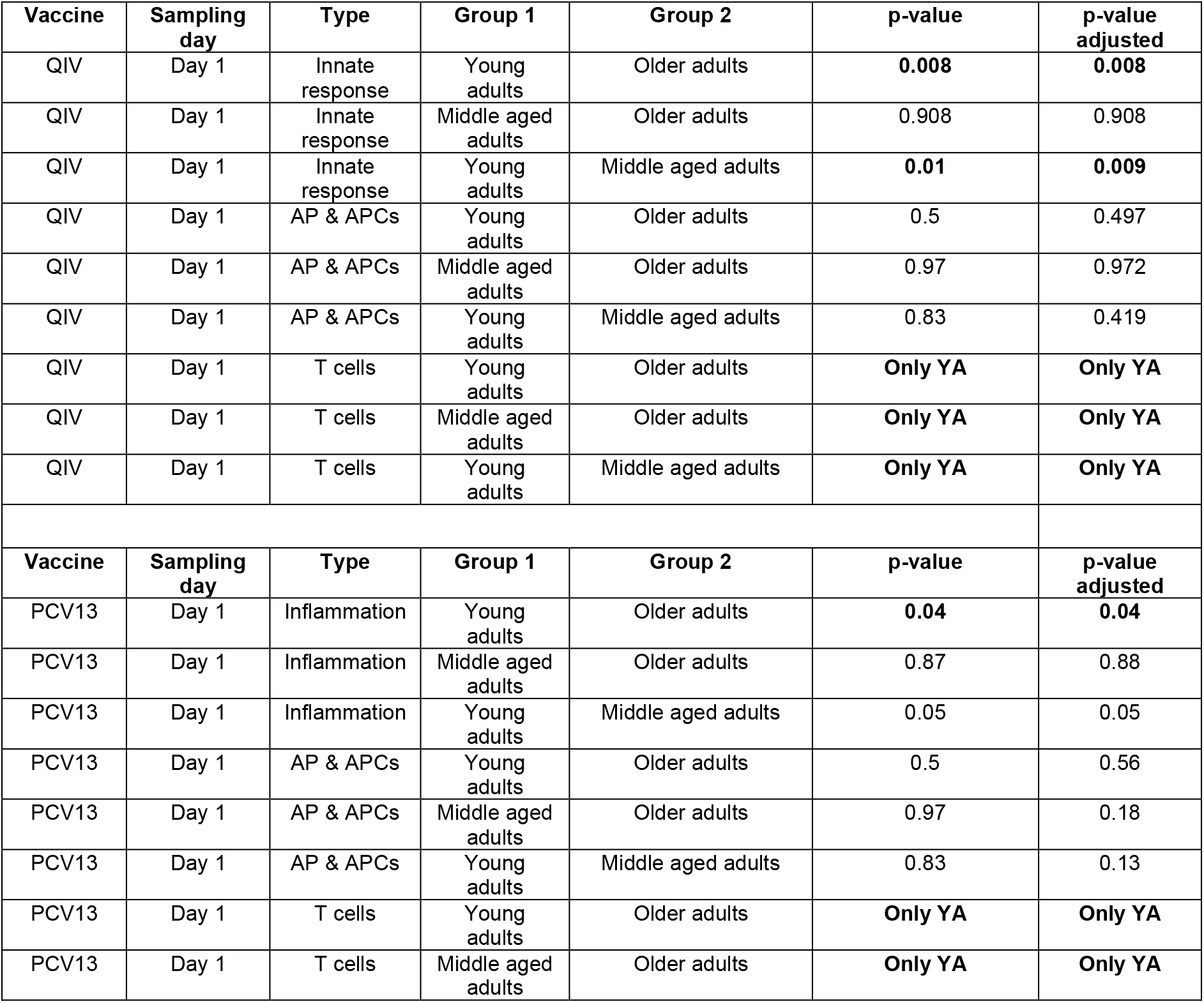
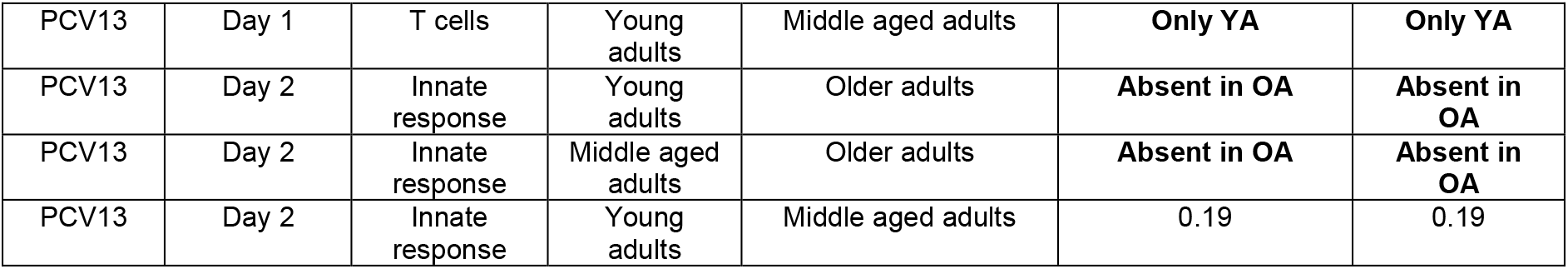
Statistical comparison (Welch’s two sample t-test) of super-module scores among the three age groups for the two vaccines. Significant differences are highlighted in bold.

**Supplementary Table 2:** Statistical comparison (Welch’s two sample t-test) of super-module scores among the two sexes for the two vaccines. Significant differences are highlighted in bold. Adjusted p-values are derived using Benjamini-Hochberg adjustment method

| Vaccine | Sampling day | Type | Group 1 | Group 2 | p-value | p-value adjusted |
| --- | --- | --- | --- | --- | --- | --- |
| QIV | Day 1 | Innate response | Male | Female | **0.001** | **0.001** |
| QIV | Day 1 | APCs | Male | Female | 0.24 | 0.25 |
| QIV | Day 1 | Inflammation | Male | Female | **0.03** | **0.03** |

**Supplementary Table 3:** Statistical comparison (Welch’s two sample t-test) of the clinical and demographic parameters within the two frailty categories. Parameters with significant differences are highlighted in bold

| Variables | Age corrected – non-frail | Age corrected - frail |
| --- | --- | --- |
| Number of participants | 44 | 33 |
| Age - mean [min-max] | 67 [51 – 89] | 65 [51 – 89] |
| Sex - % male | 47.7% | 42.4% |
| BMI - mean [min-max] | 27.4[18.3– 29.7] | 27.3[19.8– 40.6] |
| **Frailty index** – median [min – max] | 0.07[ 0.01 – 0.16] | 0.21 [0.13 – 0.53] |
| **Number of medications** – median [min – max] | 0 [0 – 10] | 4 [ 0 – 11] |
| Responder category QIV– median [min – max] | 2 [1 – 4] | 3 [1 – 4] |
| Responder category PCV13 – median [min – max] | 2 [1 – 4] | 2 [1 – 4] |

**Supplementary Table 4:** Statistical comparison (Welch’s two-sample t-test) of super-module scores among the two frailty categories for the two vaccines. Significant differences are highlighted in bold. Adjusted p-values are derived using Benjamini-Hochberg adjustment method

| Vaccine | Sampling day | Type | Group 1 | Group 2 | p-value | p-value adjusted |
| --- | --- | --- | --- | --- | --- | --- |
| QIV | Day 1 | Innate response | Frail | Non-Frail | **0.002** | **0.002** |
| QIV | Day 1 | AP & APCs | Frail | Non-Frail | 0.187 | 0.187 |
| Vaccine | Sampling day | Type | Group 1 | Group 2 | p-value | p-value adjusted |
| PCV13 | Day 1 | T-cells | Frail | Non-Frail | **Only in Non-frail** | **Only in Non-frail** |
| PCV13 | Day 1 | Inflammation | Frail | Non-Frail | **0.01** | **0.01** |
| PCV13 | Day 1 | AP & APCs | Frail | Non-Frail | 0.06 | 0.06 |

**Supplementary Table 5:** Statistical comparison (Welch’s two-sample t-test) of super-module scores among the two responder categories for the two vaccines. Significant differences are highlighted in bold. Adjusted p-values are derived using Benjamini-Hochberg adjustment method

| Vaccine | Sampling day | Type | Group 1 | Group 2 | p-value | p-value adjusted |
| --- | --- | --- | --- | --- | --- | --- |
| QIV | Day 1 | Innate response | Responder 1 | Responder 4 | **0.03** | **0.03** |
| QIV | Day 2 | Innate response | Responder 1 | Responder 4 | **only in responder 4** | **only in responder 4** |
| QIV | Day 1 | AP & APCs | Responder 1 | Responder 4 | 0.26 | 0.26 |
| Vaccine | Sampling day | Type | Group 1 | Group 2 | p-value | p-value adjusted |
| PCV13 | Day 1 | Inflammation | Responder 1 | Responder 4 | 0.17 | 0.17 |
| PCV13 | Day 2 | Inflammation | Responder 1 | Responder 4 | **only in responder 1** | **only in responder 1** |
| PCV13 | Day 1 | AP & APCs | Responder 1 | Responder 4 | 0.05 | 0.05 |

**Supplementary Table 6:** Cluster-specific expression patterns of two gene sets that are the main drivers of the transcriptome clusters. Table depicts the demographic parameters enriched for gene markers and the gene function notation

| Cluster information | Males and Older adults |  |
| --- | --- | --- |
| Highly expressed in C1 as compared to C2 and C3 | Gene name | Description |
|  | DEFA1 | Neutrophil defensins family |
|  | DEFA1B | Neutrophil defensins family |
|  | DEFA3 | Neutrophil defensins family |
|  | CEACAM8 | Response to bacterial DNA (Singer *et al.*, 2014 (57)) |
|  | LTF | Immunomodulatory molecule (Kruzel *et al.*, 2017 (58)) |
| Cluster information | Females and Older+Middle-aged adults |  |
| Highly expressed in C3 as compared to C1 and C2 | Gene name | Description |
|  | IFI44 | Interferon induced protein |
|  | IFI44L | Interferon induced protein |
|  | IFIT1 | Interferon induced protein |
|  | HERC5 | Positive regulator of innate antiviral response |
|  | CMPK2 | Immunomodulatory molecule |
|  | OAS3 | Interferon induced antiviral enzyme |

**Supplementary Table 7:** Statistical comparison (Welch’s two-sample t-test) of super-module scores among the three transcriptome clusters for the two vaccines. Significant differences are highlighted in bold. Adjusted p-values are derived using Benjamini-Hochberg adjustment method

| Vaccine | Sampling day | Type | Group 1 | Group 2 | p-value | p-value adjusted |
| --- | --- | --- | --- | --- | --- | --- |
| QIV | Day 1 | Innate response | Cluster 1 | Cluster 2 | **0.04** | **0.04** |
| QIV | Day 1 | Innate response | Cluster 2 | Cluster 3 | **0.0004** | **0.0004** |
| QIV | Day 1 | Innate response | Cluster 1 | Cluster 3 | **0.01** | **0.01** |
| QIV | Day 1 | AP & APCs | Cluster 1 | Cluster 2 | 0.38 | 0.38 |
| QIV | Day 1 | AP & APCs | Cluster 2 | Cluster 3 | No APC for Cluster 3 | No APC for Cluster 3 |
| QIV | Day 1 | AP & APCs | Cluster 1 | Cluster 3 | No APC for Cluster 3 | No APC for Cluster 3 |
| Vaccine | Sampling day | Type | Group 1 | Group 2 | p-value | p-value adjusted |
| PCV13 | Day 1 | Inflammation | Cluster 1 | Cluster 2 | 0.3 | 0.29 |
| PCV13 | Day 1 | Inflammation | Cluster 2 | Cluster 3 | **0.04** | **0.04** |
| PCV13 | Day 1 | Inflammation | Cluster 1 | Cluster 3 | **0.004** | **0.003** |
| PCV13 | Day 1 | AP & APCs | Cluster 1 | Cluster 2 | 0.33 | 0.33 |
| PCV13 | Day 1 | AP & APCs | Cluster 2 | Cluster 3 | 0.2 | 0.2 |
| PCV13 | Day 1 | AP & APCs | Cluster 1 | Cluster 3 | **0.04** | **0.04** |

**Supplementary Table 8:** Key gene markers and their association with specific immune processes. Here, UniprotKB Keyword and Gene Ontology (GO) indicate association by annotations of the respective protein entries on UniProt database.

| Day 1 - QIV - Innate and interferon response |  |
| --- | --- |
| Gene name | Source |
| GBP5 | UniprotKB Keyword |
| GBP1 | UniprotKB Keyword |
| FCGR1A | UniprotKB Keyword |
| ETV7 | Froggatt *et al.*, 2021 (59) |
| Day 1 - PCV13- Inflammation |  |
| Gene name | Source |
| SLPI | Mulligan *et al.*, 2000 (60) |
| S100A12 | UniprotKB Keyword |
| PROK2 | Watson *et al.*, 2011 (61) |
| CASP5 | Eckhart *et al.*, 2024 (62) |
| Day 7 - PCV13- Humoral/Antibody response |  |
| Gene name | Source |
| TXNDC5 | Tawfik *et al.*, 2024 (63) |
| TNFRSF17 | UniprotKB Keyword |
| JCHAIN | Gene Ontology (GO) |
| IGLL5 | Gene Ontology (GO) |

**Supplementary Table 9:**
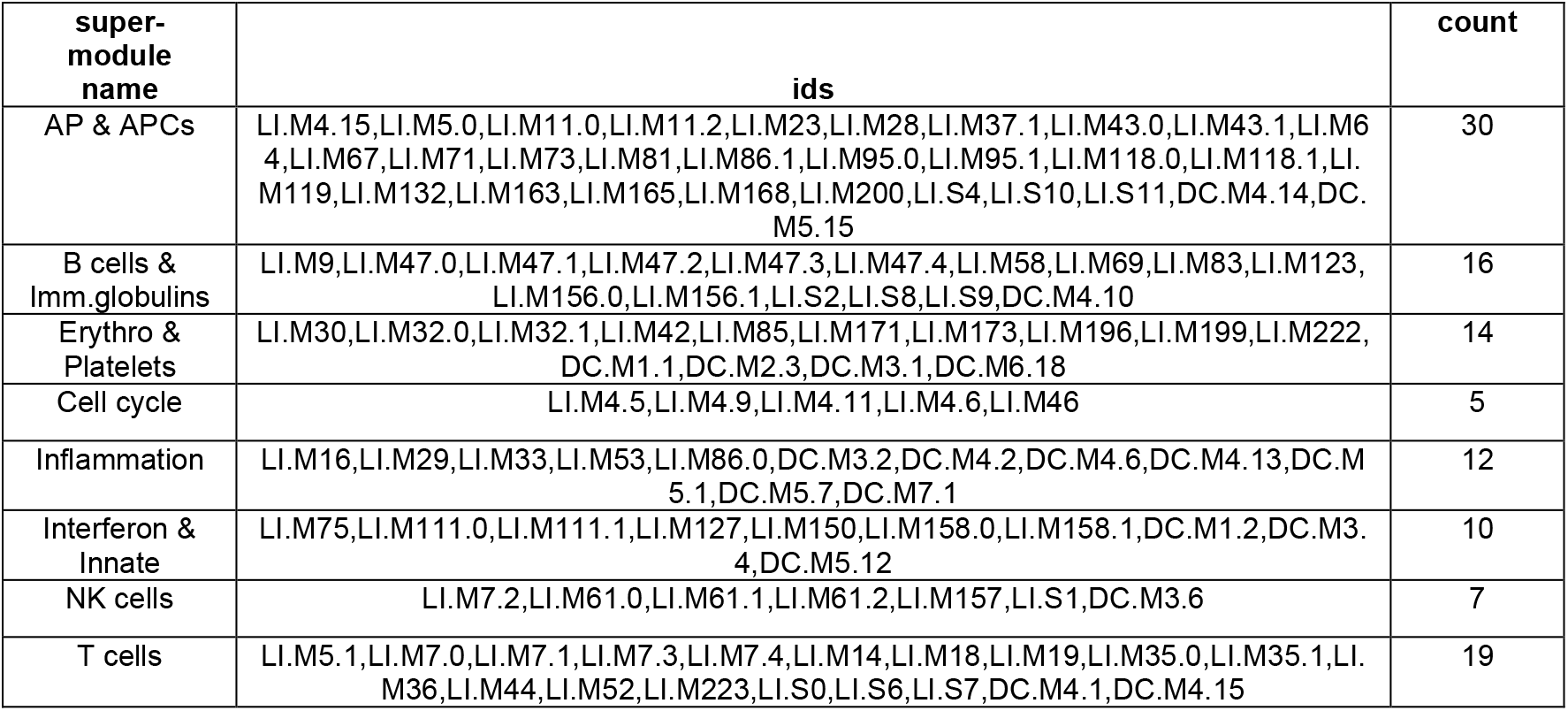
Super-module categories and the respective member IDs. Additionally, the total number of members per super-module category have been depicted

**Supplementary Table 10:**
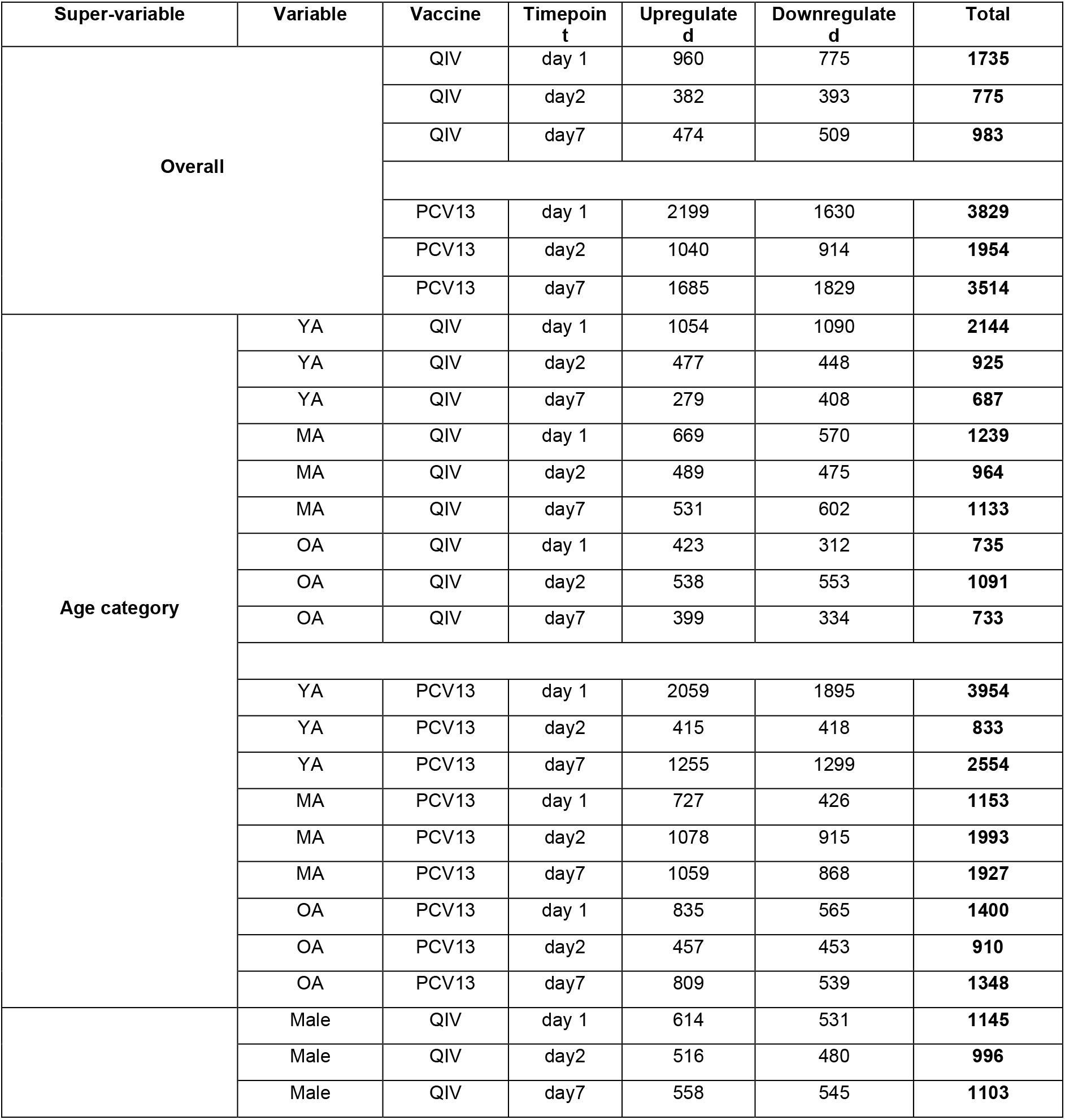
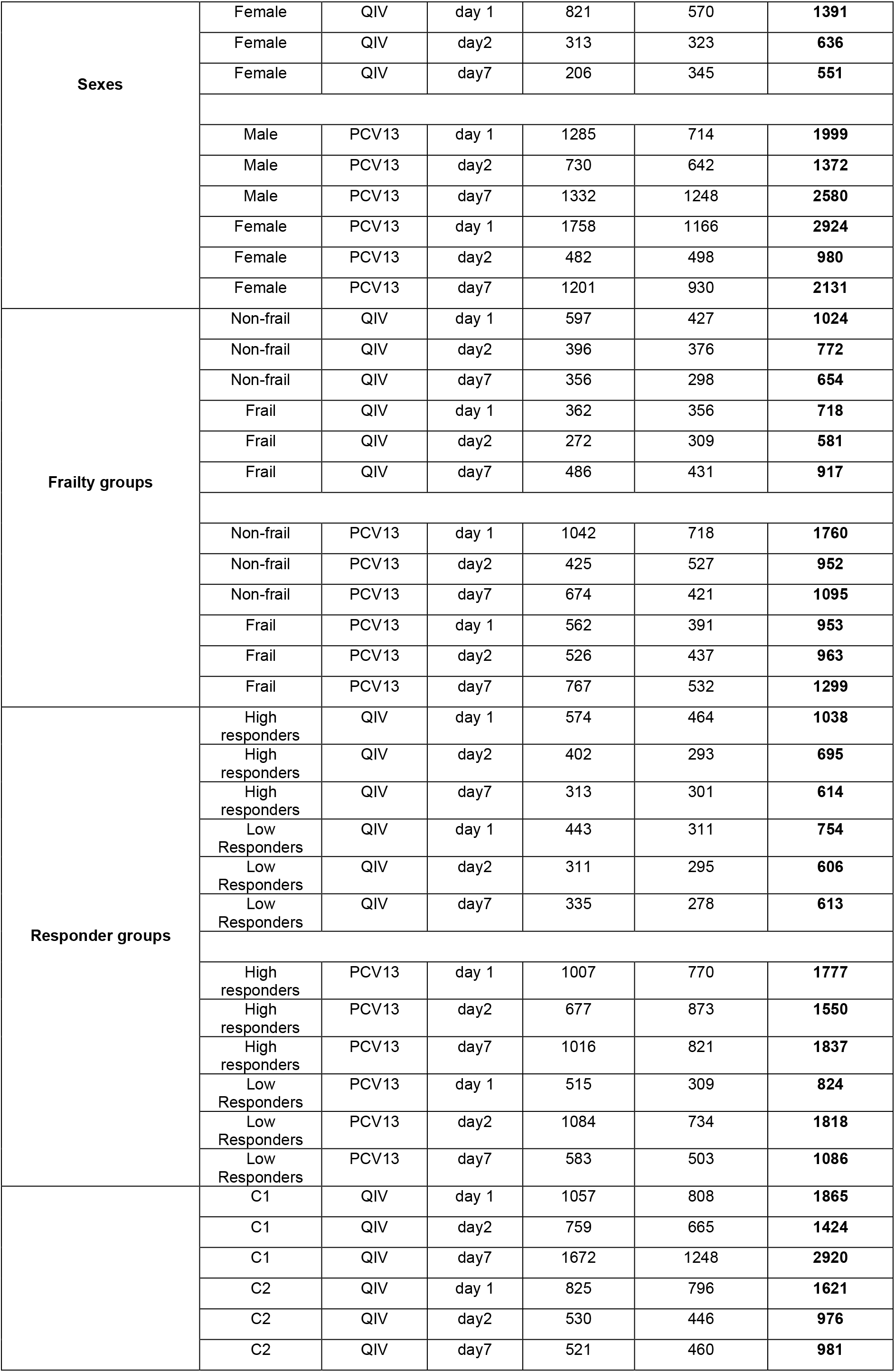
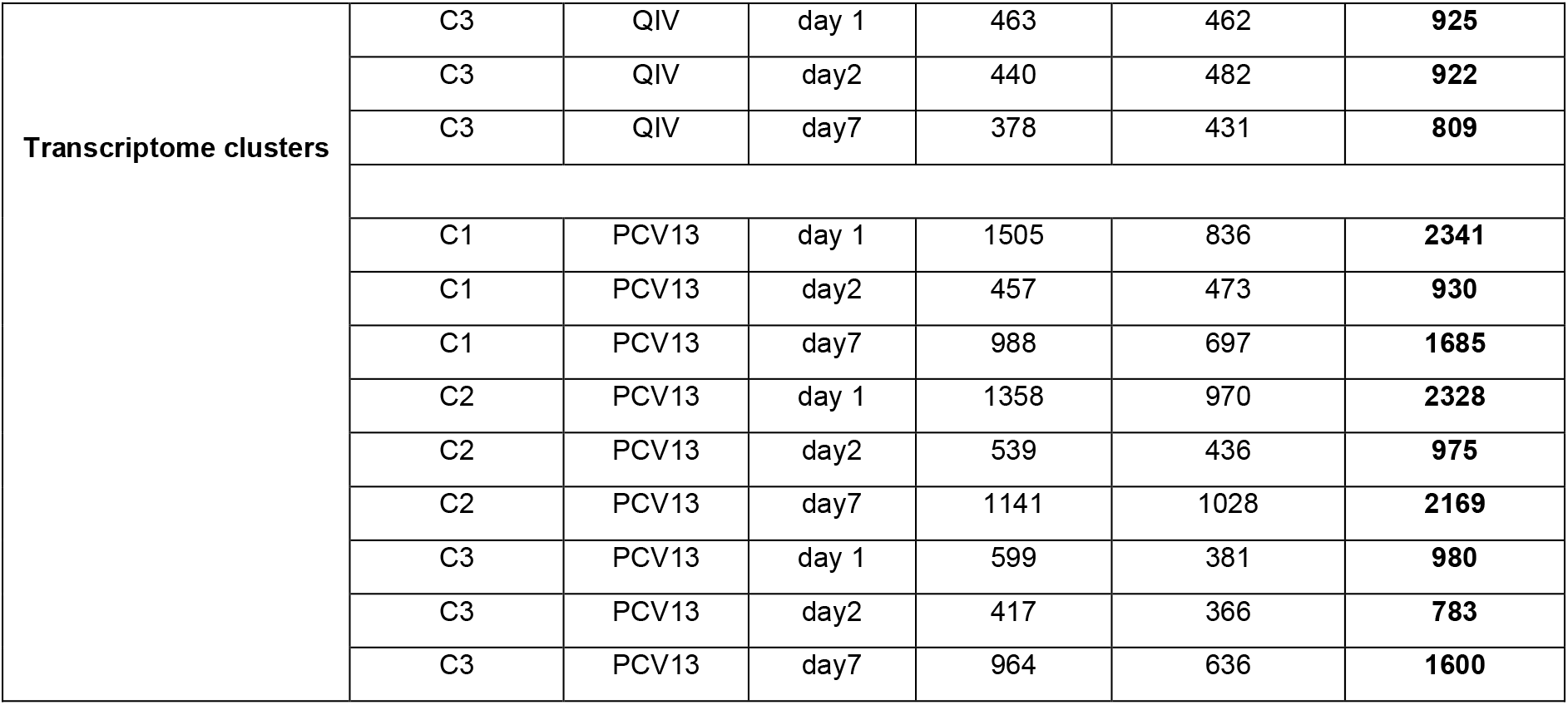
The number of differentially expressed genes (DEGs, p-value < 0.05) per comparison across the two vaccines. For every comparison, the number of upregulated and downregulated genes, along with the total DEGs (upregulated + downregulated) is depicted

**Supplementary Table 11:** BTM definitions as per Li *et al*., (2014) (*24*) and Chaussabel *et al*., (2008) (*23*) of the high-scoring modules (>3.5) across all comparisons

| Super-variable | Variable | Vaccine | Timepoint | BTM ID | BTM Name |
| --- | --- | --- | --- | --- | --- |
| Overall |  | QIV | day 1 | LI.M127 | Type I interferon response |
|  |  | QIV | day 1 | LI.M111.1 | Viral sensing & immunity; IRF2 targets network (II) |
|  |  | QIV | day 1 | DC.M3.4 | Interferon |
|  |  | QIV | day 1 | DC.M1.2 | Interferon |
|  |  | PCV13 | day 1 | DC.M3.2 | Inflammation |
|  |  | PCV13 | day 1 | LI.M16 | TLR and inflammatory signalling |
|  |  | PCV13 | day 1 | DC.M4.2 | Inflammation |
|  |  | PCV13 | day 1 | LI.M64 | Enriched in activated dendritic cells/monocytes |
|  |  | PCV13 | day 1 | LI.M37.1 | Enriched in neutrophils (I) |
|  |  | PCV13 | day 1 | LI.M132 | Recruitment of neutrophils |
|  |  | PCV13 | day 7 | LI.M156.1 | Plasma cells, immunoglobulins |
| Age category | YA | QIV | day 1 | DC.M1.2 | Interferon |
|  | YA | QIV | day 1 | DC.M3.4 | Interferon |
|  | YA | QIV | day 1 | LI.M150 | Innate antiviral response |
|  | YA | QIV | day 1 | LI.M127 | Type I interferon response |
|  | YA | QIV | day 1 | LI.M111.1 | Viral sensing & immunity; IRF2 targets network (II) |
|  | YA | PCV13 | day 1 | DC.M3.2 | Inflammation |
|  | YA | PCV13 | day 1 | LI.M16 | TLR and inflammatory signalling |
|  | YA | PCV13 | day 1 | DC.M4.2 | Inflammation |
|  | YA | PCV13 | day 1 | LI.M7.4 | T cell activation (III) |
|  | YA | PCV13 | day 1 | LI.M37.1 | Enriched in neutrophils (I) |
|  | YA | PCV13 | day 7 | LI.M156.1 | Plasma cells, immunoglobulins |
|  | MA | PCV13 | day 7 | LI.M156.1 | Plasma cells, immunoglobulins |
|  | OA | PCV13 | day 7 | LI.M156.1 | Plasma cells, immunoglobulins |
| **Sexes** | Female | QIV | day 1 | LI.M150 | Innate antiviral response |
|  | Female | QIV | day 1 | DC.M1.2 | Interferon |
|  | Female | QIV | day 1 | DC.M3.4 | Interferon |
|  | Female | QIV | day 1 | LI.M127 | Type I interferon response |
|  | Female | QIV | day 1 | LI.M111.1 | Viral sensing & immunity; IRF2 targets network (II) |
|  | Female | PCV13 | day 1 | DC.M3.2 | Inflammation |
|  | Female | PCV13 | day 1 | LI.M132 | Recruitment of neutrophils |
|  | Female | PCV13 | day 7 | LI.M156.1 | Plasma cells, immunoglobulins |
|  | Male | PCV13 | day 7 | LI.M156.1 | Plasma cells, immunoglobulins |
| **Frailty categories** | Non-frail | PCV13 | day 7 | LI.M156.1 | Plasma cells, immunoglobulins |
|  | Frail | PCV13 | day 7 | LI.M156.1 | Plasma cells, immunoglobulins |
| **Responder categories** | High Responders | QIV | day 1 | LI.M111.1 | Viral sensing & immunity; IRF2 targets network (II) |
|  | High Responders | QIV | day 1 | LI.M127 | Type I interferon response |
|  | High Responders | QIV | day 1 | DC.M3.4 | Interferon |
|  | High Responders | PCV13 | day 7 | LI.M156.1 | Plasma cells, immunoglobulins |
|  | Low Responders | PCV13 | day 7 | LI.M156.1 | Plasma cells, immunoglobulins |
| **Transcriptome clusters** | C2 | QIV | day 1 | LI.M150 | Innate antiviral response |
|  | C2 | QIV | day 1 | LI.M111.1 | Viral sensing & immunity; IRF2 targets network (II) |
|  | C2 | QIV | day 1 | DC.M1.2 | Interferon |
|  | C2 | QIV | day 1 | DC.M3.4 | Interferon |
|  | C2 | QIV | day 1 | LI.M127 | Type I interferon response |
|  | C1 | PCV13 | day 7 | LI.M156.1 | Plasma cells, immunoglobulins |
|  | C2 | PCV13 | day 7 | LI.M156.1 | Plasma cells, immunoglobulins |
|  | C3 | PCV13 | day 7 | LI.M156.1 | Plasma cells, immunoglobulins |
| **Decay categories** | High Responders | QIV | day 1 | LI.M111.1 | Viral sensing & immunity; IRF2 targets network (II) |
|  | High Responders | QIV | day 1 | LI.M127 | Type I interferon response |
|  | High Responders | QIV | day 1 | DC.M3.4 | Interferon |
|  | High Responders | QIV | day 1 | DC.M1.2 | Interferon |
|  | High Responders | PCV13 | day 7 | LI.M156.1 | Plasma cells, immunoglobulins |
|  | Low Responders | PCV13 | day 7 | LI.M156.1 | Plasma cells, immunoglobulins |

**Supplementary Table 12:** PCV-13 strain-specific IgG concentrations (in µg/mL) at pre-vaccination and 28-day post-vaccination timepoints in the three age categories. The concentrations are provided as median [min-max]

| Serotypes | Timepoint | Young-adults | Middle-aged-adults | Older-adults |
| --- | --- | --- | --- | --- |
| Serotype 1 | Pre-vaccination | 0.3 [0.005 – 12.4] | 0.2 [0.01 – 8.9] | 0.1 [0.005 – 16.3] |
|  | 28-day post-vaccination | 11.8 [0.06 – 438.6] | 6.6 [0.1 – 201.5] | 4.4 [0.2 – 255.4] |
| Serotype 3 | Pre-vaccination | 0.4 [0.03 – 3.9] | 0.4 [0.02 – 20.9] | 0.2 [0.01 – 22.8] |
|  | 28-day post-vaccination | 2.7 [0.5 – 74.7] | 1.7 [0.2 – 29.5] | 1.3 [0.08 – 67.3] |
| Serotype 4 | Pre-vaccination | 0.1 [0.002 – 6.0] | 0.1 [0.002 – 2.5] | 0.1 [0.001 – 1.6] |
|  | 28-day post-vaccination | 6.0 [0.6 – 23.2] | 3.1 [0.03 – 131.0] | 1.8 [0.02 – 41.9] |
| Serotype 5 | Pre-vaccination | 0.7 [0.06 – 6.2] | 0.5 [0.02 – 8.4] | 0.3 [0.01 – 15.4] |
|  | 28-day post-vaccination | 18.4 [1.5 – 413.1] | 13.6 [0.3 – 601.0] | 9.5 [0.25 – 456.2] |
| Serotype 6A | Pre-vaccination | 0.3 [0.002 – 7.7] | 0.4 [0.008 – 16.7] | 0.3 [0.002 – 60.9] |
|  | 28-day post-vaccination | 17.1 [0.6 – 1206.0] | 7.7 [0.05 – 371.5] | 5.8 [0.05 – 509.0] |
| Serotype 6B | Pre-vaccination | 0.2 [0.004 – 3.7] | 0.3 [0.004 – 6.1] | 0.2 [0.004 – 7.1] |
|  | 28-day post-vaccination | 11.4 [0.4 – 240.9] | 3.9 [0.08 – 191.6] | 2.8 [0.03 – 301.2] |
| Serotype 7F | Pre-vaccination | 0.3 [0.01 – 13.7] | 0.6 [0.01 – 10.8] | 0.4 [0.006 – 7.2] |
|  | 28-day post-vaccination | 16.1 [0.7 – 576.4] | 14.7 [0.4 – 283.9] | 14.0 [0.4 – 272.4] |
| Serotype 9V | Pre-vaccination | 0.2 [0.01 – 1.9] | 0.3 [0.008 – 2.5] | 0.1 [0.008 – 43.1] |
|  | 28-day post-vaccination | 4.6 [0.3 – 79.7] | 5.7 [0.1 – 108.0] | 2.9 [0.05 – 92.0] |
| Serotype 14 | Pre-vaccination | 0.2 [0.01 – 10.0] | 0.8 [0.02 – 15.4] | 0.3 [0.01 – 24.9] |
|  | 28-day post-vaccination | 6.2 [0.03 – 76.9] | 8.2 [0.02 – 135.6] | 7.2 [0.02 – 204.1] |
| Serotype 18C | Pre-vaccination | 0.4 [0.02 – 2.5] | 0.4 [0.01 – 11.2] | 0.4 [0.02 – 21.9] |
|  | 28-day post-vaccination | 12.6 [0.4 – 75.3] | 10.9 [0.07 – 156.3] | 15.9 [0.2 – 937.2] |
| Serotype 19A | Pre-vaccination | 0.5 [0.05 – 16.5] | 0.5 [0.02 – 19.8] | 0.8 [0.006 – 16.7] |
|  | 28-day post-vaccination | 10.8 [0.9 – 288.2] | 12.5 [0.06 – 169.2] | 13.4 [0.1 – 406.6] |
| Serotype 19F | Pre-vaccination | 0.4 [0.006 – 11.0] | 0.6 [0.006 – 26.0] | 0.5 [0.02 – 12.6] |
|  | 28-day post-vaccination | 9.7 [0.2 – 408.9] | 7.9 [0.21 – 102.7] | 5.6 [0.1 – 183.3] |
| Serotype 23F | Pre-vaccination | 0.2 [0.01 – 6.2] | 0.3 [0.003 – 6.8] | 0.3 [0.003 – 22.6] |
|  | 28-day post-vaccination | 18.9 [0.2 – 343.2] | 5.5 [0.007 – 376.0] | 3.6 [0.01 – 239.1] |

